# Splenic CD169^+^Tim4^+^ Marginal Metallophilic Macrophages Are Essential for Wound Healing After Myocardial Infarction

**DOI:** 10.1101/2024.08.09.24311769

**Authors:** Mohamed Ameen Ismahil, Guihua Zhou, Min Gao, Shyam S. Bansal, Bindiya Patel, Nita Limdi, Min Xie, Sergey Antipenko, Gregg Rokosh, Tariq Hamid, Sumanth D. Prabhu

## Abstract

Fidelity of wound healing after myocardial infarction (MI) is an important determinant of subsequent adverse cardiac remodeling and failure. Macrophages derived from infiltrating Ly6C^hi^ blood monocytes are a key component of this healing response; however, the importance of other macrophage populations is unclear. Here, using a variety of in vivo murine models and orthogonal approaches, including surgical myocardial infarction, splenectomy, parabiosis, cell adoptive transfer, lineage tracing and cell tracking, RNA sequencing, and functional characterization, we establish in mice an essential role for splenic CD169^+^Tim4^+^ marginal metallophilic macrophages (MMMs) in post-MI wound healing. Splenic CD169^+^Tim4^+^ MMMs circulate in blood as Ly6C^low^ cells expressing macrophage markers and help populate CD169^+^Tim4^+^CCR2^−^LYVE1^low^ macrophages in the naïve heart. After acute MI, splenic MMMs augment phagocytosis, CCR3 and CCR4 expression, and robustly mobilize to the heart, resulting in marked expansion of cardiac CD169^+^Tim4^+^LyVE1^low^ macrophages with an immunomodulatory and pro-resolving gene signature. These macrophages are obligatory for apoptotic neutrophil clearance, suppression of inflammation, and induction of a reparative macrophage phenotype in the infarcted heart. Splenic MMMs are both necessary and sufficient for post-MI wound healing, and limit late pathological remodeling. Liver X receptor-α agonist-induced expansion of the splenic marginal zone and MMMs during acute MI alleviates inflammation and improves short- and long-term cardiac remodeling. Finally, humans with acute ST-elevation MI also exhibit expansion of circulating CD169^+^Tim4^+^ macrophages. We conclude that splenic CD169^+^Tim4^+^ MMMs are required for pro-resolving and reparative responses after MI and can be manipulated for therapeutic benefit to limit long-term heart failure.

**CLINICAL PERSPECTIVE:** *What is new?:*

- We establish for the first time that metallophilic marginal macrophages (MMMs) from the spleen, expressing the markers CD169 and Tim4, circulate in blood and traffic to the heart to help maintain the CD169^+^Tim4^+^CCR2^−^LYVE1^low^ macrophage population in the heart.
- After acute myocardial infarction, splenic MMMs augment cardiac trafficking in response to chemotactic signals, resulting in expansion of CD169^+^Tim4^+^ macrophages in the heart that play an essential role in post-MI efferocytosis, wound healing and repair while limiting longer term adverse cardiac remodeling.
- Analogous to mice, humans also exhibit circulating CD169^+^Tim4^+^ macrophages in the blood that expand after acute ST segment elevation MI.

*What are the clinical implications?:*

- This study highlights the importance of the cardiosplenic axis in acute MI, and the splenic marginal zone, in determining the course and outcome of post-MI LV remodeling.
- Pharmacological expansion of splenic marginal zone macrophages alleviated post-MI adverse LV remodeling and inflammation, suggesting that splenic modulation is a potential translational therapeutic approach for limiting post-MI inflammation and improving heart repair.

## INTRODUCTION

Myocardial infarction (MI) triggers an orchestrated wound healing response initially comprised of intense inflammation and clearance of dead cells, subsequently followed by inflammation resolution, wound healing, and scar formation.^1^ While this biphasic response is required for effective tissue repair, an inflammatory response that is excessively vigorous, or fails to resolve in a timely manner, can lead to deleterious LV remodeling and long-term heart failure (HF).^1^ Innate immune cells are critical for this process. The initial inflammatory phase is dominated early by infiltrating neutrophils, and subsequently by Ly6C^hi^CX3CR1^low^ monocytes derived from the spleen and bone marrow, and pro-inflammatory macrophages.^2–6^ In the healing phase, reparative Ly6C^low^ macrophages are generated locally and proliferate in the heart. These cells are also derived from the initial surge of Ly6C^hi^ monocytes, via a process dependent on the nuclear hormone receptor Nr4a1.^2^ The specific triggers responsible for the switch from pro-inflammatory to reparative cells in the infarcted heart are not fully defined, but may in part relate to the phagocytic clearance of dead cells, including apoptotic cardiomyocytes and neutrophils,^7, 8^ that induces a pro-resolving phenotype in macrophages.^1^

While the role of Ly6C^hi^ monocytes and Ly6C^hi^ monocyte-derived macrophages post-MI has been well-delineated, the potential importance of other macrophage populations remains unclear. The subcapsular red pulp of the spleen is an important reservoir of Ly6C^hi^ monocytes that mobilize to the infarcted heart.^3, 5, 9^ Beyond the red pulp, however, the spleen contains several spatially localized macrophage populations with specialized functions. Prominent among these are macrophages in the marginal zone (MZ) that surrounds the splenic white pulp.^10, 11^ The MZ is an important watershed between lymphoid tissue and blood that is characterized by continuous leukocyte transit. Along with SIGN-R1^+^MARCO^+^ MZ macrophages (MZMs), the mouse MZ harbors CD169(sialoadhesin)^+^ marginal metallophilic macrophages (MMMs) that play crucial roles in antigen capture and processing, communication with dendritic cells, T cells, and B cells, regulation of apoptotic cell clearance, and facilitation of immune tolerance.^10–14^ CD169^+^ MMMs coordinate host immune responses that impact both inflammation and immunoregulation.^15^

The phosphatidylserine receptor Tim4 (T cell immunoglobulin- and mucin-domain-containing molecule 4) allows for the recognition and phagocytosis of apoptotic cells,^16^ and, along with LYVE1, has been proposed as a specific marker of resident tissue macrophages.^17–19^ Tim4 further identifies an important tissue CD169^+^ macrophage subset capable of suppressing local immune responses.^20^ As proper healing post-MI necessitates both inflammation resolution and efficient apoptotic cell clearance, we posited that CD169^+^Tim4^+^ MMMs from the spleen may play a central role in these events. Nonetheless, whether splenic CD169^+^Tim4^+^ MMMs mobilize to the heart during acute MI is unknown. Moreover, the pathophysiological role of splenic MMMs in post-MI repair, and the subsequent development of adverse LV remodeling and HF, is unexplored. Accordingly, we tested the hypothesis that splenic CD169^+^Tim4^+^ MMMs infiltrate the acutely infarcted heart and play an essential role in inflammation resolution and wound healing post-MI.

## METHODS

### Mouse models

All studies were performed in compliance with the National Research Council’s Guide for the Care and Use of Laboratory Animals (revised 2011). Male CD45.1 (#002014) and CD45.2 (#000664) C57BL/6 male mice, and macrophage Fas-induced apoptosis (MaFIA) mice expressing an EGFP and conditional suicide construct transgene downstream of the *c-fms* promoter^21^ (#005070) were purchased from Jackson Laboratories. CX3CR1^CreERT2^; Rosa26^tdTomato^ mice were generated by breeding CX3CR1^CreERT2^ (#020940) and Rosa26^tdTomato^ mice (#007914), which have a *loxP*-flanked STOP cassette preventing transcription of a CAG promoter-driven red fluorescent tdTomato (TdT) gene. *Cre*-specific expression removes the STOP sequence allowing TdT expression.^22^ Cre expression was induced in 7-week-old mice upon treatment subcutaneously with a pulse of 10 mg 4-hydroxytamoxifen (Sigma-Aldrich) dissolved in 200 ml corn oil. Controls were littermates with the loxP-flanked allele but lacking Cre recombinase. CD169-DTR *Tg* mice (C57BL/6 background) with human DTR cDNA introduced into the CD169 gene,^13^ thereby allowing transient cell depletion with DT,^23^ were obtained from the RIKEN BioResource Center (Japan). We used heterozygous (CD169^DTR/+^) mice bred by crossing CD169^DTR/DTR^ mice with C57BL/6 mice. Catchup (Ly6G^Cre-tdTomato^) and Catchup^IVM-red^ (Ly6G^Cre-tdTomato^;Rosa26[Rtom]) mice were obtained from Dr. Matthias Gunzer and used as neutrophil reporter lines.^24^ The Institutional Animal Care and Use Committees at the University of Alabama at Birmingham (UAB) and Washington University in St. Louis gave local approval for these studies. Mouse numbers are provided in the figure legends.

### Murine surgical procedures

Coronary ligation and sham surgery were performed under anesthesia with 1-1.5% inhaled isoflurane and mechanical ventilation as previously described.^25, 26^ Animal mortality in the first 24 h post-surgery was excluded from Kaplan-Meier survival assessment. Survival splenectomy was performed as previously described.^26, 27^ For mouse parabiosis, we used the protocol of Kamran et al.^28^ Briefly, two male mice (one CD45.1 and other CD45.2, or one CD169^DTR^ and the other MaFIA) of similar size, co-housed for at least 2 w, were anesthetized with 1.0–1.5% isoflurane. The opposing flanks of each mouse were shaved and disinfected with topical iodine and alcohol. The skin of each mouse was incised from elbow to knee and interconnected with a continuous suture. Buprenorphine (0.1 mg/kg s.c.) was given for analgesia postoperatively. Four weeks after surgery, coronary ligation or sham operation was performed in the host mouse after establishment of donor blood chimerism with flow cytometry. In some host and donor mice, splenectomy was performed 2 w prior to parabiosis surgery.

### Echocardiography

Mouse echocardiography was performed under 1.5% inhaled isoflurane anesthesia (with 100% supplemental O_2_) using a VisualSonics Vevo770 High-Resolution System and 30 MHz RMV707B scanhead, and a heated, bench-mounted adjustable rail system as previously described.^26, 27, 29^ Data analysis and assessment of LV size and systolic function were performed using VisualSonics software as previously described.^29^

### Mononuclear cell isolation

Mononuclear cells were isolated from blood, heart, spleen, bone marrow as previously described,^26^ with slight modification of cardiac immune cell isolation as follows. Briefly, whole hearts were flushed with PBS to remove blood and minced into 2 mm pieces with a single-edged blade. Tissue was digested in RPMI media with collagenase-2 (1 mg/mL, Worthington), trypsin (1 mg/mL, Invitrogen), and DNase I (10 ug/mL) at 37°C for 45 min with gentle agitation. Cell suspensions were filtered through a 100 μm cell strainer (BD Biosciences) and incubated with 2 mM EDTA in PBS for 5 min at 37°C. Cell pellets were collected and washed with PBS. Cells were fixed with 1% paraformaldehyde and stored in staining buffer (eBioscience) at 4°C.

### Flow cytometry and cell labeling

Cell suspensions were incubated with anti-mouse CD16/32 (clone 93, BioLegend) for 10 min at 4°C to block Fcγ receptors, and then stained for 60 minutes in staining buffer with anti-mouse fluorochrome-conjugated antibodies in panel appropriate combinations for specific experiments as follows: Ly6C-PE-Cy7 (HK1.4, eBioscience), Ly6C-PE Vio770 (1G7.G10, Miltenyi Biotec), CD45.1-FITC (A20, Miltenyi Biotec), CD45.2-PE (104, BD Biosciences), CD169-PE (REA197, Miltenyi Biotec; and 3D6.112, BioLegend), CD11b-Alexa Fluor 700 (M1/70, eBioscience), Tim4-Alexa Fluor 647 (RMT4-54, BioLegend), Gr1/Ly6G-eFluor 450 (RB6-8C5, eBioscience), F4/80-PerCP-Cy5.5 (BM8, eBioscience), MARCO-FITC (ED31, Novus Biologicals), CD45-600 Super Bright (30-F11, eBioscience), CD45-PE-Cy7 (30-F11, BD Biosciences), LYVE1 Alexa Fluor 488 (ALY7, eBioscience), CD45-605 NC (30-F11, eBioscience), CD54(ICAM-1)-PE (eBioKat-1, eBioscience), MHCII (I-A/I-E)-APC-eFluor780 (M5/114.15.2, eBioscience), TGFβ1-APC (TW7-16B4, BioLegend), CD54-FITC (YN1/1.7.4, BioLegend), CD169-PerCP/Cy5.5 (3D6.112, BioLegend), CD169-PE (3D6.112, BioLegend), CD64-Brilliant Violet 605 (X54-5/7.1, BioLegend), CD206-FITC (MR5D3, AbD Serotech), CD45-eVolve 605 (30-F11, eBioscience), CD206-Alexa Fluor 647 (MR5D3, Bio-Rad), IL17A-Alexa Fluor 700 (TC11-18H10.1, BioLegend), CD34-Alexa Fluor 647 (SA376A4, BioLegend), CD117(c-kit)-PE/Cy7 (ACK2, BioLegend), NOS2-TRITC (N-20, Santa Cruz Biotechnology), and CD16/32-PE (93, BioLegend, without pre-Fcγ receptor blocking).

For specific assessment of bone marrow granulocyte-monocyte precursors (GMPs), we used a pacific blue-conjugated lineage (Lin) antibody cocktail (BioLegend) comprised of anti-mouse CD3 (17A2); Ly-6G/Ly-6C (RB6-8C5); CD11b (M1/70); CD45R/B220 (RA3-6B2); TER-119/erythroid cells (Ter-119). For intracellular staining of IL-4, IL10 and TGF-β, cells were permeabilized with PBS containing 0.5% v/v tween-20 prior to antibody labeling for 60 minutes. Apoptotic neutrophils were identified by staining with Annexin V-FITC (Life Technologies). Flow cytometry was performed with the BD LSR II (BD Biosciences), and data acquisition/analysis performed with FlowJo 10.6.1 software. Fluorescence compensation control and unstained samples were used to accurately identify cells within multicolor-stained samples. tSNE analysis was run using the FlowJo dimensionality reduction algorithm plugin (iterations=2000, perplexity=20 and θ=0.5). Samples were randomly downsampled to 3000 events per sample and then concatenated prior to dimensionality reduction analysis. tSNE heat maps indicate fluorescent intensity of the relevant cell surface marker, with scales generated from low to high expression.

Splenic red pulp, marginal zone, and marginal zone metallophilic macrophages were identified as CD45^+^CD11b^low^F4/80^hi^, CD45^+^CD11b^low^F4/80^low^MARCO^+^, and CD45^+^CD11b^low^ F4/80^low^CD169^+^ cells, respectively.^30–32^ Bone marrow granulocyte-macrophage progenitors (GMPs) were defined as Lin^−^c-kit^+^CD34^+^ CD16/32^+^ cells.^33^

### Assessment of CD169^+^Tim4^+^ macrophage phagocytosis *in vivo*

One day post-MI or sham operation, mice were intravenously administered 10 mg/kg Texas Red conjugated-Zymosan-A (*S. cerevisiae*) bioparticles (Molecular Probes) and sacrificed 3 h later for mononuclear cell isolation. Bioparticle^+^CD169^+^Tim4^+^ cells in the blood, heart, and spleen were analyzed by flow cytometry. In some mice, 100 μg anti-Tim4 neutralizing antibody (G-6, Santa Cruz Biotechnology) was given i.v. at the time of MI.

### Complete blood count

Cheek vein blood was collected into sodium citrate tubes. Individual blood cell counts were measured with a Hemavet 950 system (Drew Scientific).

### Myocardial gene expression

Total RNA from MI border zone or sham LV (≈10 mg) was extracted using Trizol, and gene expression was measured using RT-PCR as described previously.^25, 26, 29^ *<u>Table S2</u>* lists the forward and reverse primer sequences used for the genes of interest.

### Immunostaining and microscopy

Formalin-fixed, paraffin-embedded MI hearts were sectioned at 5 μm thickness, deparaffinized, and rehydrated. Masson trichrome staining was used to evaluate tissue fibrosis as previously described,^25, 26, 29, 34^ Images were obtained with bright field light microscopy (Nikon Eclipse TE2000E) and fibrosis (blue staining) quantified in an automated manner using Metamorph software v6.3r5 (Molecular Devices).

Immunofluorescent staining was performed using tissue embedded in optimal cutting temperature compound (Tissue Tek) and cryopreserved at −80^°^C.^29, 34^ Tissue sections (∼8-10 μm) were thawed at room temperature, and fixed with 1% paraformaldehyde for 20 min. Sections were blocked and incubated at 4^°^C for 2 h (as indicated) with anti-mouse CD169-PE (Miltenyi Biotec), Tim4-Alexa Fluor 647 (BioLegend), MARCO-FITC (AbD Serotec), F4/80 (Invitrogen, with rat IgG1-efluor 660 secondary antibody), Ly6G-PE (eBioscience), CD206-FITC (Bio-Rad), iNOS-PE (eBioscience), MMP-9 (Millipore-Sigma, with anti-rabbit-Alexa Fluor 555 secondary antibody from Life Technology), IL-10-APC (eBioscience), CD45.1-FITC (MACS), and secondary antibodies conjugated with Alexa Fluor 488, 555, or 647 (Invitrogen) if needed. After washing with PBS, sections were incubated with DAPI (Molecular Probes) for nuclear staining. Images were obtained using a Zeiss LSM710 confocal microscope at 63X magnification, and Z-stack images were acquired according to standard protocols.

### Splenic CD169^+^ MMM depletion

Genetic depletion was performed in CD169^DTR/+^ mice upon i.p. injection with 10 μg/kg DT (List Biological, Lot #15043A1) in 100 μL PBS. For pharmacological depletion, mice were injected i.v. with 4 mg/kg of clodronate-loaded liposomes (Clo-Lip) in 100 μL suspension solution (Encapsula Nanosciences). This low dose of clo-lip selectively depletes macrophages in the splenic marginal zone.^32, 35^ PBS-loaded liposomes were used as a control.

### Splenic CD169^+^Tim4^+^ MMM adoptive transfer

Live splenocytes from naïve CD45.1 donor mice were antibody labeled with CD169-PE and Tim4-Alexa Fluor 647. CD169^+^Tim4^+^ cells were sorted with a BD FACSAria II sorter (BD Biosciences) to a purity of >90%. CD169^+^Tim4^+^ cells (∼1X10^6^) were adoptively transferred via tail vein injection (100 μL PBS) into recipient splenectomized CD45.2 mice with MI as indicated in specific protocols.

### Sample Preparation for bulk RNA Sequencing

Live mononuclear cells isolated from 1 d post MI heart were stained with SYTOX, and CD45, CD64, Tim4, CD169 and LYVE1 antibodies. SYTOX^−^CD45^+^CD64^+^Tim4^+^CD169^+^LYVE1^high^ or LYVE1^low^ macrophages were sorted on a BD FACS Aria flow cytometer. For spleen and blood, cells were stained and sorted based on SYTOX^−^CD45^+^Ly6C^low^CD169^+^ and SYTOX^−^CD45^+^Ly6C^low^CD169^-^ antibodies. Using SMART-Seq v4 Ultra Low Input RNA Kit (Takara Bio USA, Inc.) 250 cells were collected in lysis buffer at room temperature, pelleted and processed for cDNA library preparation according to the kit protocol. Briefly, mRNA in cell lysates were reverse transcribed and cDNA was prepared for amplification by template switching and extension. cDNA was amplified with 21 PCR cycles and purified for Illumina Nextera DNA library construction and sequencing. STAR (version 2.5.3a) was used to align the raw RNA Seq FASTQ reads to the reference genome from Gencode.^36^ Cufflinks was used on the STAR-aligned reads to assemble transcripts, estimate their abundances, and test for differential expression and regulation.^37, 38^ Cuffmerge was used to merge the assembled transcripts to a reference annotation and track Cufflinks transcripts across multiple experiments.

### RNA sequencing data analysis

The raw fastq files were processed using the default settings of each analysis tool except as specified below. Standard Illumina adaptors were used to trim reads with TrimGalore (version 0.4.5). The ENCODE options for standard long RNA-seq were utilized for mapping using STAR (version 2.5.3), as defined in the STAR manual^36^, and Ensembl Mus musculus genome (GRCm38) was the reference sequence. Quality of libraries was assessed by overall mapping rate. The htseq-count script in the HTSeq package (version 0.7.2) was used to calculate transcript abundance estimates. The R software environment (version 3.6.1) with BioConductor was used for statistical analysis. Differential expression analysis was performed using R package DESeq2 (version 1.18.1).^39^ The statistical analysis of differential expression (DE) between conditions using gene count tables generated by htseq-count pipeline. Gene counts were filtered for genes catalogued as protein coding. These filtered files were imported in DESeq2 using the DESeqDataSetFromHTSeqCount function. The gene counts were normalized and log_2_-transformed followed by application of the linear model to the comparison between conditions. The statistics were reported including log fold change (logFC), p-value, and adjusted p-value (padj) corrected for multiple hypothesis testing with the Benjamini-Hochberg procedure. The thresholds of padj < 0.05 were used to obtain significant differentially expressed genes. The rlog function from the DESeq2 package was utilized for variance stabilization and was applied to all samples. The principal component plots were performed using the top 500 differentially expressed genes after rlog transformation. MA plots were generated using R package ggplot2 (version 3.3.3). The DEGs (adjusted p value q<0.05) genes in samples of pairwise comparisons were visualized with heatmaps using the R package Complex Heatmap (version 2.2.0).

### Gene set enrichment analysis (GSEA)

GSEA was performed to explore whether the identified sets of genes showed significant differences between the two groups. To prepare data for gene set enrichment analysis, results of differential gene expression analysis were ranked by signed p-value. The gene sets KEGG pathways and Gene Ontology Biological Process (GO-BP) in MsigDB were used to perform the enrichment analysis. A p-value quantifying the likelihood that a given gene set displays the observed level of enrichment for differentially expressed genes was calculated using R package fgsea (version 1.12.0) with 10,000 permutations. Gene set enrichment p-values of Normalized Enrichment Scores (NES) were corrected with the Benjamini-Hochberg procedure. The GSEA results were visualized by bar plots of NES values using R package ggplot2 (version 3.3.3). The colors of blue or red bars based on the negative or positive NES. The length of bar was directly proportional to the NES value generated from the enrichment of each pathway for each comparison. Barcode plots were generated with the plotEnrichment function from the fgsea package.

### Single cell RNAseq analysis

Mouse peripheral blood underwent erythrocyte lysis and leukocytes were centrifuged and collected immediately. Cells were stained with 2 μL DAPI (564907, BD Pharmingen) and 2 μL Draq5 (62251, Thermo Scientific) for 5 min. Cells were diluted to 4 mL for sorting (FACSMelody, BD Biosciences), and viability was verified by ethidium homodimer (EtHD) staining (E1169, Thermo Fisher Scientific). Cells were processed for scRNAseq using the 10X fixed sample preparation kit (1000414, 10X Genomics). Cells were pelleted and resuspended in 1 mL fixation buffer and maintained at 4^°^C for 24 h. Cells were then collected at 850*g* at 5 min at RT and resuspended in 1 mL chilled quenching buffer. Cell concentration was determined by EtHD staining (Countess, Thermo Fisher Scientific) and then stored at −80 ^°^C after addition of 100 μL enhancer and glycerol (10% final concentration). Cells were processed by the Washington University McDonnell Genome Institute (MGI) for scRNAseq using the 10X genomics 16-plex format (10X Genomics CDG000527 Rev E). Cell barcoding on the 10X Chromium X, library construction, amplification and sequencing were performed, and scRNAseq FASTQ files obtained from the MGI were processed using 10X genomics cell ranger pipeline for sequence quality, demultiplexing, and alignment. Processed FASTQ files were exported as H5 files and uploaded into BioTuring for clustering and cell type classification.

### Data and code availability

RNA sequencing data has been deposited in GEO, accession number: GSE210798. All data and code are available upon request.

### Human studies

Peripheral blood was collected from human subjects upon initial presentation to UAB Hospital with acute ST-elevation MI under the auspices of a UAB IRB-approved protocol (#X151201004). Blood was also collected from control subjects (with coronary artery disease but without acute coronary syndrome) immediately prior to undergoing elective percutaneous coronary intervention (PCI) under the auspices of UAB IRB protocol #X130807012. Informed consent was obtained from all subjects, whose clinical characteristics are listed in *<u>Table S1</u>*. Blood cells were collected and processed for flow cytometry as above. The following anti-human fluorochrome-conjugated antibodies were used: CD45-BV605 (H130), CD64-PerCP/ Cy5.5 (10.1), Tim4-PE/Cy7 (9F4) (BioLegend); CD14-FITC (BD Biosciences), HLA-DR-Alexa Fluor 700 (LN3, eBiosciences), CD169-APC (7-239, Invitrogen).

### Imaging flow cytometry

Sorted human blood CD14^+^HLA-DR^+^CD169^+^ cells were imaged with an Amnis ImageStream®^X^ Mk II Imaging Flow Cytometer, using a 100x oil-immersed objective lens, numerical aperture 1.45. Images were processed using IDEAS Version 6.1 software.

### Statistical analysis

All results are mean±SD. Analyses were performed using GraphPad Prism version 7.03 software. Group variances were compared using the Brown−Forsythe test, whereas normality was assessed using the D’Agostino–Pearson test. Two-group statistical comparisons were performed using an unpaired *t* test with equal or unequal variance for normally distributed variables or Mann-Whitney *U* test for non-normal distribution. For comparisons of more than 2 groups, experimental datasets were first assessed for normality. For normally distributed data, a 1- or 2-way analysis of variance was performed, with Bonferroni, Tukey’s, or Dunnett’s T3 post-test to adjust for multiple comparisons. For single cell RNAseq, gene expression differences were assessed using t-test with false discovery rate (FDR) correction (BioTuring BBrowserX software, version 3.5.26). If a non-normal distribution was observed, the data were logarithmically transformed prior to assessment. Specific approaches are in the figure legends. A p value of <0.05 was considered statistically significant.

## RESULTS

### Splenic CD169^+^Tim4^+^ MMMs circulate in blood and populate the heart

Spleens were harvested from naïve wild-type (WT) C57BL/6 mice. Immunostaining identified splenic CD169^+^Tim4^+^ macrophages mainly confined to the MZ (<u>*Figure 1A*</u>). Using the gating strategy shown in *<u>Figure S1A</u>*<u>, s</u>plenic MMMs (CD45^+^Ly6C^−^CD11b^low^F4/80^low^CD169^+^) were identified by flow cytometry; ∼55% of CD169^+^ MMMs were Tim4^+^ (*<u>Figure 1A</u>*). We next evaluated for circulating CD169^+^Tim4^+^ cells. Using the FACS strategy in *<u>Figure S1B</u>*, we identified Ly6G^−^ Ly6C^+^CD169^+^ Tim4^+^ cells in the blood, which were primarily (>95%) in the Ly6C^low^ compartment (*<u>Figure S1C</u>*). May Grünwald-Giemsa staining of FACS-sorted cells revealed a distinct morphology (cell projections) and smaller size than circulating Ly6C^hi^ monocytes (*<u>Figure S1D</u>*).^5^ Further analysis of Ly6C^low^ cells revealed a minor subpopulation expressing CD169 *(Figure 1B)*. Principal component analysis (PCA) of the top 500 differentially expressed genes (DEGs) from RNAseq of sorted cells revealed segregation of CD169^+^ and CD169^−^ subsets. Transcriptomic profiling revealed 334 DEGs (q<0.05) and discrete clustering of the two Ly6C^low^ subsets, suggesting functionally distinct populations. CD169^+^Tim4^+^ cells comprised ∼12-13% of the blood Ly6C^low^ cell population, and ∼90% of these cells expressed CD64, considered a core macrophage marker,^40–42^ and MHCII (*<u>Figure 1B</u>*), suggesting a macrophage identity. Notably, mice 4 w after splenectomy (Spx) exhibited profound (∼80%) reductions in circulating Ly6C^low^CD169^+^Tim4^+^ cells. Spx mice also exhibited mild leukocytosis, neutrophilia, anemia, and thrombocytosis (*<u>Figure S2A</u>*), but no changes in circulating B cells, CD4^+^ and CD8^+^ T cells, and Ly6C^hi^ monocytes, and comparable cardiac size and function (*<u>Figure S2B-C</u>*).

**Figure 1.**
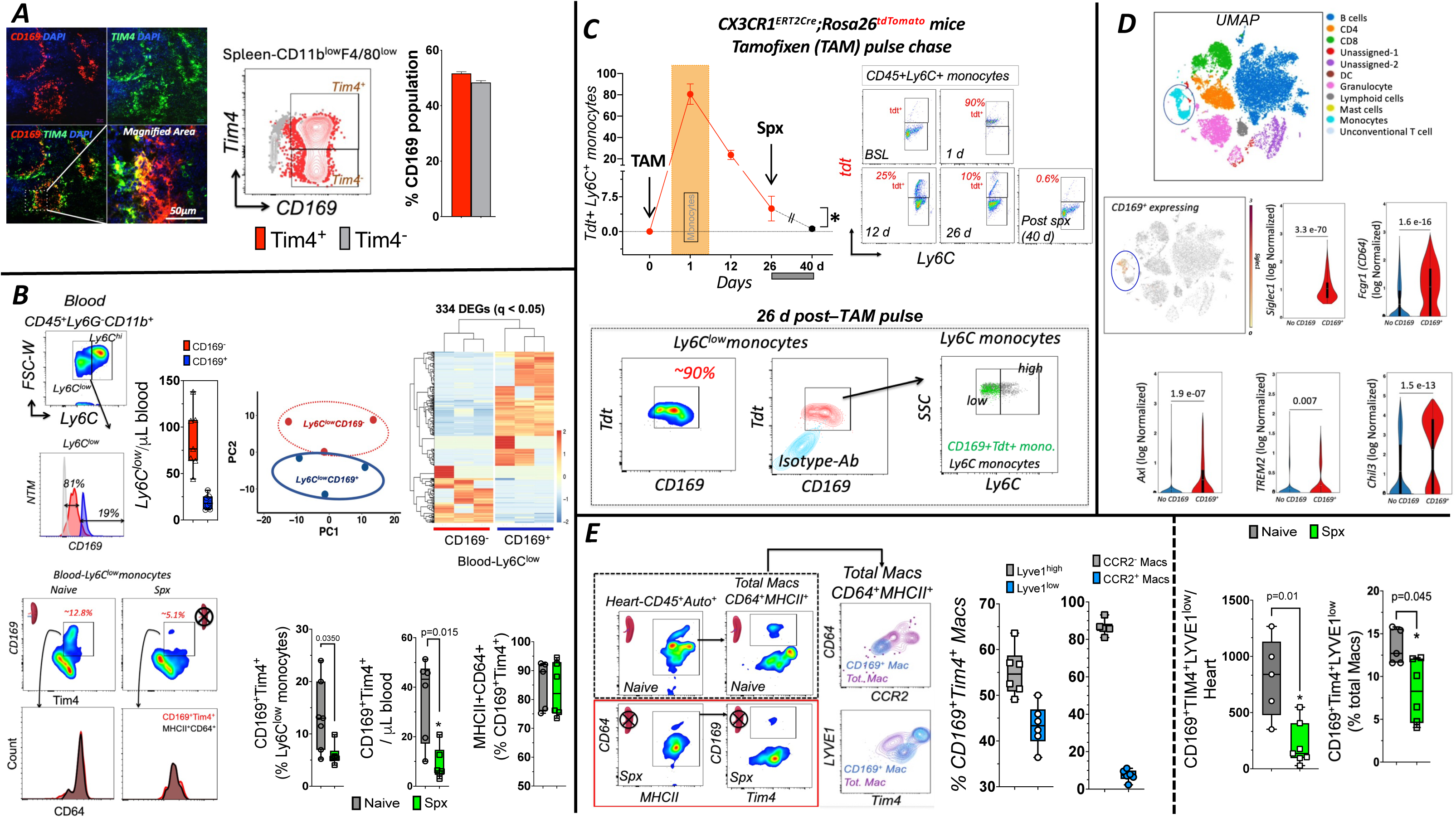
Splenic CD169^+^Tim4^+^ marginal metallophilic macrophages (MMMs) circulate as Ly6C^low^ leukocytes and populate the naïve heart. ***A*.** *Left*, Representative immunostaining of macrophages in the splenic marginal zone expressing CD169 (red) and Tim4 (green); DAPI nuclear staining (blue). *Right*, FACS contour plots for CD169 and Tim4 expression in splenic CD45^+^CD11b^low^F4/80^low^ cells and quantitation of Tim4 expression in CD169^+^ MMMs; n=4. ***B*.** *Top Left*, FACS pseudocolor plots of circulating Ly6C monocytes with histograms identifying CD169^+^ and CD169^−^ populations within Ly6C^low^ monocytes, and corresponding cell quantitation; NTM, normalized to mode. *Top Right*, principal component analysis of top 500 differentially expressed genes (DEGs) from bulk RNAseq of sorted Ly6C^low^CD169^−^ and Ly6C^low^CD169^+^ cells, and heat maps with dendrograms for 334 DEGs (q<0.05) between the same sub-populations. *Bottom*, FACS pseudocolor plots of circulating Ly6C^low^CD169^+^Tim4^+^ macrophages in naïve and spx mice blood with flow histograms identifying CD64 and MHCII surface expression (in black), together with quantitation of frequency or absolute number of the populations shown; n=5-7, statistics: unpaired *t* test. ***C.*** *Top,* Representative blood FACS dot plots and quantitation of tdTomato (Tdt) expression in blood Ly6C^+^ monocytes from CX3CR1^CreERT2^; Rosa26^tdTomato^ mice after a tamoxifen (TAM) pulse to induce Cre recombination. Shown are data from several time points after TAM and 14 d after splenectomy (Spx) performed at 26 d post-TAM; *p<0.05, n=3, statistics: unpaired t-test. *Bottom*, representative FACS plots showing CD169 expression in Tdt+ cells 26 d post-TAM pulse and overlay of the CD169^+^Tdt^+^ cells (green) on blood Ly6C^+^ monocytes. ***D*.** *Top*, UMAP plots derived from single cell RNA sequencing of blood leukocytes from 3 naïve mice (∼400K total cells, 11 identified cell clusters), heat map demonstrating CD169 (*Siglec1*) expression primarily in the monocyte cluster, and quantitative expression of select macrophage genes in cells with and without CD169 expression (adjusted FDR p values are shown). ***E*.** *Left,* FACS plots identifying CD169^+^Tim4^+^ cardiac macrophages in intact and Spx mice, overlay of these macrophages on contour plots of CCR2 and LYVE1 expression (in intact mice), and quantitation of overall LYVE1 and CCR2 expression (n=6). *Right*, quantitation of frequency and number of cardiac CD169^+^Tim4^+^LYVE1^low^ macrophages naïve and Spx C57BL/6 mice; n=5-7/group, statistics: unpaired *t* test.

To definitively establish macrophage identity of blood CD169^+^Tim4^+^ cells, we performed lineage tracing in Cx3cr1^Cre-ERT2^;Rosa26^tdTomato^ mice in which a tamoxifen pulse labels all Cx3cr1-expressing cells (monocytes, macrophages, dendritic cells [DCs]) with tdTomato. Given shorter lifespan, monocytes and DCs no longer express tdTomato by 3 weeks as new cells replace them, whereas macrophage tdTomato is retained.^43^ As illustrated in *<u>Figure 1C</u>*, 1 d after tamoxifen, ∼80% of Ly6C^+^ monocytes expressed tdTomato. At 26 d after tamoxifen, ∼6% of total Ly6C^+^ blood cells remained td-Tomato^+^ supporting macrophage identity. Most of these td-Tomato^+^ cells expressed CD169 and were primarily Ly6C^low^. Notably, repeat analysis 2 weeks after splenectomy in a subset of these mice revealed near total disappearance of circulating Ly6C^+^tdT^+^ cells, consistent with circulating CD169^+^ macrophages of splenic origin. Moreover, single cell RNAseq analysis of blood leukocytes from naïve mice (*<u>Figure 1D</u>*) revealed CD169 (*Siglec1*) expression in a minority (14%) of cells identified as monocytes. As compared to monocytes without *Siglec1*, *Siglec1* expressing cells exhibited significantly higher transcript levels of macrophage genes *Fcgr1 (CD64), Axl, Trem2*, and *Chil3*.

We next characterized cardiac CD169^+^Tim4^+^ macrophages. Live/dead staining with 7-aminoactinomycin D (7-AAD) routinely yielded >90-95% viability of isolated heart mononuclear cells (*<u>Figure S3A</u>*). We identified CD64^+^MHCII^+^CD169^+^Tim4^+^ macrophages using gating strategy 1 in *<u>Figure S3B</u>*, and used LYVE1, a marker specific for resident macrophages,^17, 44, 45^ to delineate LYVE1^hi^ (cardiac resident^17^) and LYVE1^low^ macrophage subsets. As shown in *<u>Figure 1E</u>*, cardiac CD169^+^Tim4^+^ macrophages were principally (∼90%) CCR2^−^ but exhibited variable LYVE1 expression, with ∼40% being LYVE1^low^. Notably, Spx mice exhibited significant reduction (∼80%) of cardiac CD169^+^Tim4^+^LYVE1^low^ macrophages (*<u>Figure 1E</u>*) without significant change in the LYVE1^hi^ subset (*<u>Figure S4A</u>*). PCA of bulk RNAseq of sorted CD64^+^MHCII^+^CD169^+^Tim4^+^ cardiac macrophages revealed clear separation based on LYVE1 expression, with 987 DEGs (q<0.05) and discrete clustering of LYVE1^hi^ and LYVE1^low^ cells, suggesting distinct functional subsets (*<u>Figure S4B</u>*). Moreover, expression analysis of 54 select macrophage- and DC-associated genes^46–48^ revealed both subsets expressed macrophage marker genes, but uniformly low levels of DC-associated genes (*<u>Figure S4C</u>*). Intravascular CD45 labeling *in vivo* immediately prior to cardiac harvest in a subset of mice (*<u>Figure S5</u>*) revealed that intravascular leukocytes comprised ∼0.5% of cardiac CD169^+^Tim4^+^ cells isolated at steady-state, and <2% 24 h after MI, confirming tissue localization. Collectively, these findings support a circulating population of splenic CD169^+^Tim4^+^ MMMs that populate the steady-state heart and are primarily LYVE1^low^ cells.

### Splenic CD169^+^Tim4^+^ macrophages augment cardiac trafficking and phagocytosis acutely after MI

WT mice were evaluated 24 h after non-reperfused MI, during the inflammatory phase of repair. As compared with sham-operated mice, there was a ∼3-fold increase in blood CD169^+^Tim4^+^ macrophages 24 h post-MI (*<u>Figure 2A)</u>*. CCR3 and CCR4 expression increased in circulating CD169^+^Tim4^+^ macrophages (*<u>Figure 2B</u>*) along with upregulation of cognate chemokine ligands in border zone myocardium, including CCL5, CCL6, CCL7, CCL17, and CCL22.^49^ In contrast, Spx mice did not exhibit increases in circulating CD169^+^Tim4^+^ cells 24 h post-MI (*<u>Figure 2C</u>*). We next examined CD169^+^Tim4^+^ macrophage infiltration in the acutely infarcted heart. Here, to comport with prior studies of MMMs in the spleen, and to maintain consistency between approaches in blood and heart given dynamic innate immune cell flux acutely after MI, we used low expression of Ly6C as a central identifier of CD169^+^Tim4^+^ macrophages <u>as illustrated</u> in *<u>Figure S3B</u>*, gating strategy 2. As illustrated in *<u>Figure 2D</u>*, WT mice exhibited a robust ∼2.5-fold increase in cardiac CD169^+^Tim4^+^ macrophages 1 D after M; overall cell numbers were reduced in Spx mice and the increase after MI was abrogated. Concomitantly, the spleen in MI mice exhibited dynamic changes with significant hypotrophy and loss of marginal metallophilic macrophages, suggesting their egress from the spleen (*<u>Figure 2E</u>*). We next evaluated phagocytic activity of Ly6C^low^CD169^+^Tim4^+^ macrophages. Fluorescent bioparticles were given i.v. 3 h prior to sacrifice with determination of cell bioparticle uptake by FACS. As shown in *<u>Figure 2F</u>*, there was a significant (∼2-fold) increased uptake of bioparticles by CD169^+^Tim4^+^ cells in both blood and heart versus sham mice, establishing enhanced phagocytic capacity of these macrophages after MI. Interestingly, there was an attendant reduction of CD169^+^Tim4^+^bioparticle^+^ macrophages in the spleen, consistent with overall fewer MMMs early after MI.

**Figure 2.**
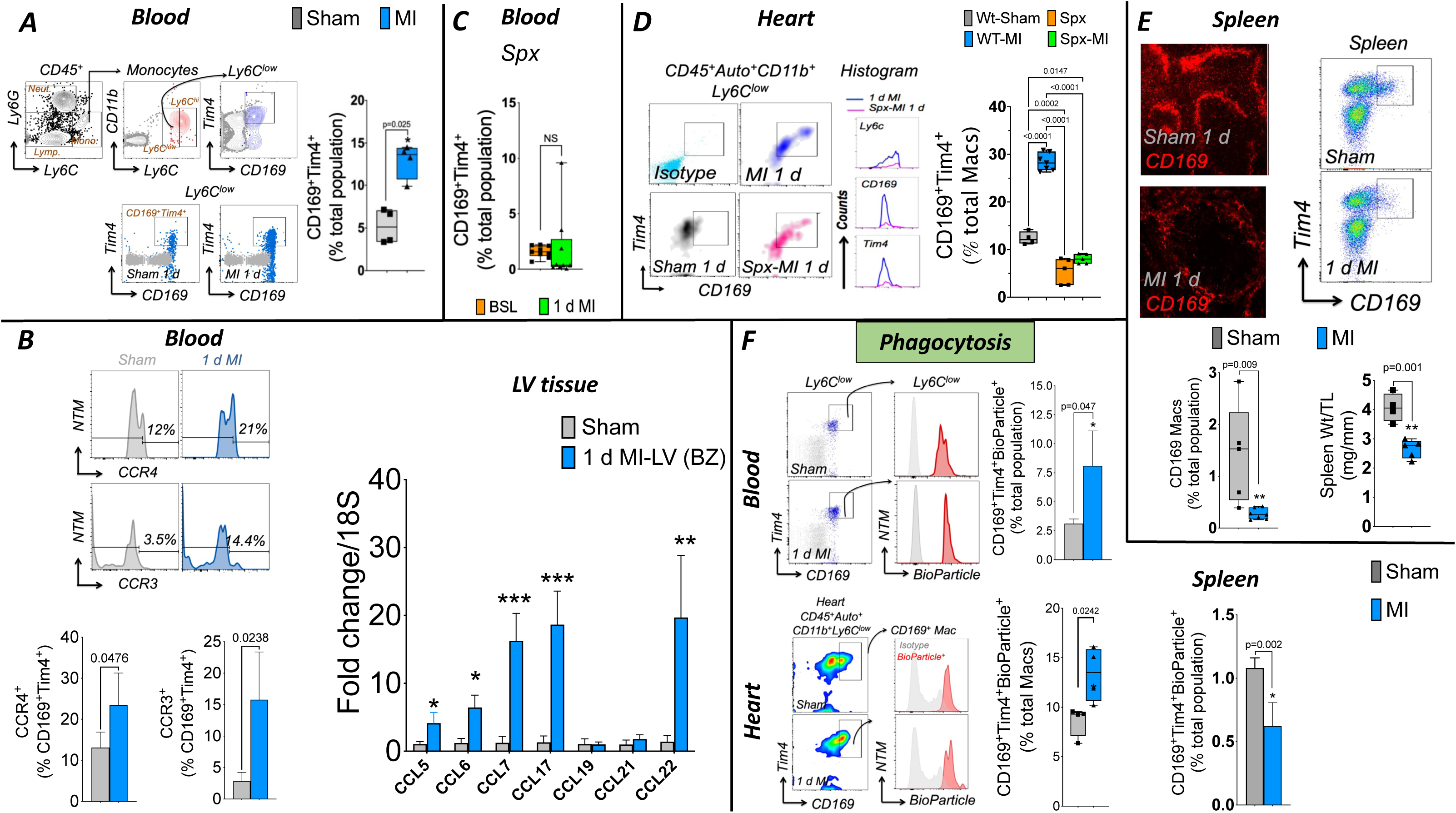
Splenic MMMs increase cardiac trafficking and phagocytosis after acute MI. ***A*.** FACS plots and group quantitation for circulating Ly6C^low^CD169^+^Tim4^+^ macrophages in sham-operated and MI wild type (WT) C57BL/6 mice 1 d post-MI; n=4/group, statistics: unpaired *t* test. ***B.*** *Left*, Flow histograms demonstrating surface expression of CCR3 and CCR4 in Ly6C^low^CD169^+^Tim4^+^ macrophages from the same groups; statistics: unpaired *t* test, NTM, normalized to mode, y-axis represents cell counts. *Right*, chemokine gene expression by RT-PCR (normalized to 18s) in the myocardial border zone (BZ) 1 d after MI or sham operation; n=4-5/group, statistics: unpaired *t* test. *p<0.05, **p<0.01, ***p<0.001 versus sham. ***C*.** *Left*, Circulating Ly6C^low^CD169^+^Tim4^+^ cell frequency prior to and 1 d after MI in Spx mice; n=9, statistics: paired *t* test. NS, not significant. ***D.*** FACS density plots, histograms, and quantitation of cardiac Ly6C^low^CD169^+^Tim4^+^ macrophages in WT and Spx mice 1 d post-MI or sham operation; n=4-7/group, statistics: unpaired *t* test. ***E***. Immunostains and FACS dot plots of splenic CD169^+^ MMMs (red) 24 h after MI or sham operation, and quantitation of MMM frequency by FACS and spleen weight (Wt); n=5-7/group, statistics: unpaired *t* test. TL, tibia length. ***F***. *Left*, FACS dot plots and histograms, and corresponding quantitation, of blood and heart Ly6C^low^CD169^+^Tim4^+^Bioparticle^+^ cells from sham and MI mice given 10 mg/kg Texas Red-conjugated bioparticles i.v. 3 h before sacrifice; n=3-4/group, statistics: unpaired *t* test for blood and non-parametric Mann-Whitney *U* test for heart (non-normal distribution). Right, quantitation of splenic BioParticle^+^ MMMs in the same experimental mouse groups; statistics: unpaired t test.

To definitively establish a splenic source for CD169^+^Tim4^+^ macrophages in the acutely infarcted heart, we used parabiosis. The circulations of CD45 isotype-mismatched mice were surgically joined, with non-reperfused MI induced 4 w later in either intact or Spx host mice (*<u>Figure S6</u>*). Blood FACS (2 d post-MI) indicated stable ∼35% donor chimerism of CD45^+^ blood leukocytes in both intact and Spx host mice (*<u>Figures S6 and 3A</u>*). We then evaluated cardiac CD169^+^ macrophages using gating strategy 2 in *<u>Figure S3</u>*. In hearts of intact host MI mice, ∼15% of CD169^+^ macrophages were of donor origin; notably, donor chimerism significantly increased by 138% in Spx host MI mice (*<u>Figure 3A</u>*), consistent with augmented sourcing from the parabiont donor spleen.

**Figure 3.**
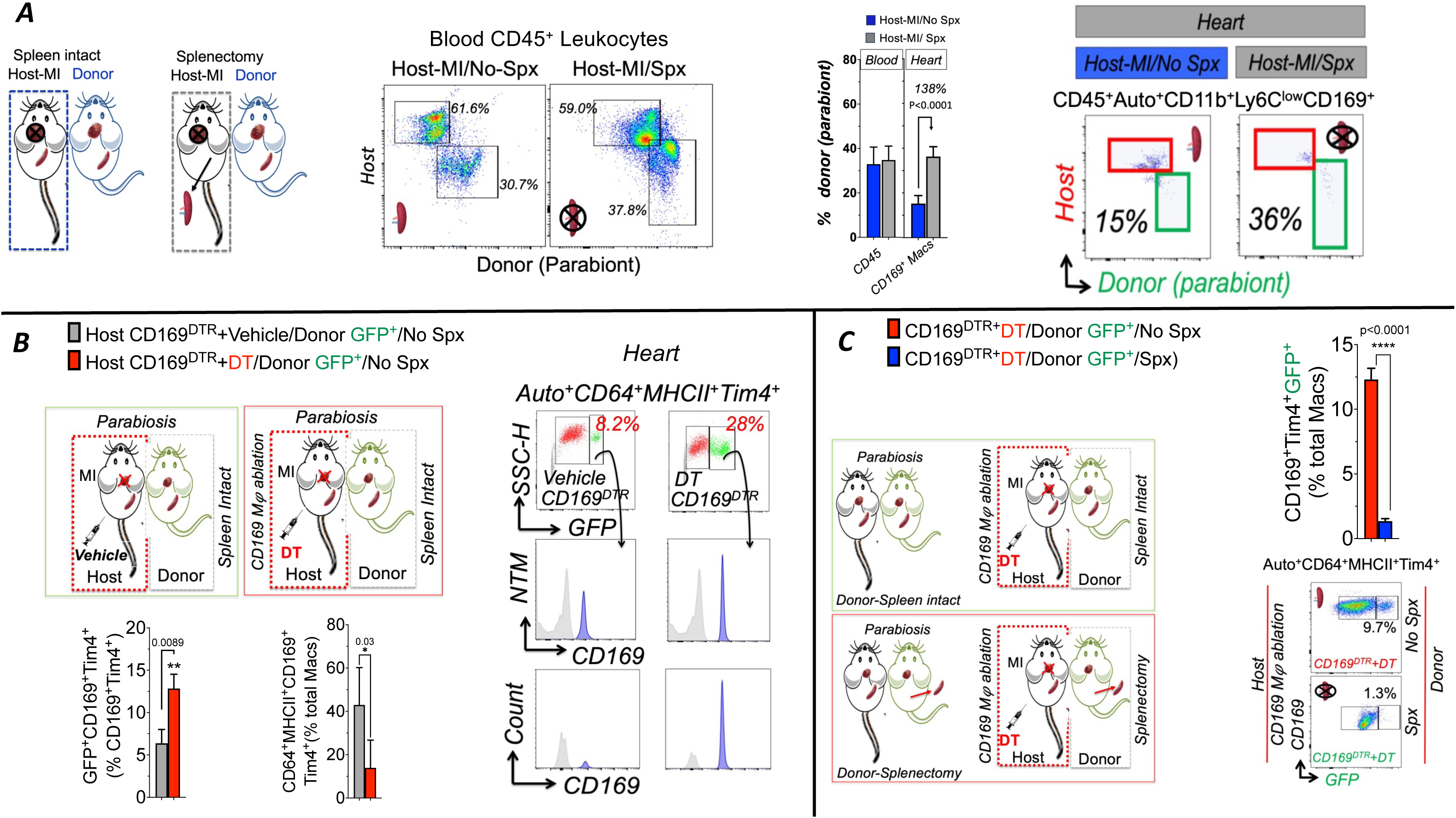
The spleen is an important source for CD169^+^Tim4^+^ macrophages in the acutely infarcted heart. ***A*** *Left*, Parabiosis schema joining CD45 isotype-mismatched host (spleen-intact or after splenectomy [Spx]) and donor parabiont mice, with MI induced in the host. *Middle and Right*, FACS plots and quantitation of donor chimerism in host mouse blood (total CD45^+^ leukocytes) and heart (Ly6C^low^CD169^+^ macrophages) in spleen-intact and Spx host mice 48 h after MI. n=4-7/group, statistics: unpaired t-test. ***B*** *Top Left*, Parabiosis schema joining CD169^DTR^ host and donor parabiont MaFIA mice, with host mice given either vehicle or diphtheria toxin (DT) at the time of MI. *Right*, Representative FACS dot plots of donor GFP^+^CD169^+^ macrophages in 48 h post MI hearts from host mice and flow histograms of CD169 expression in GFP^+^CD64^+^MHCII^+^Tim4^+^ cells delineated as NTM or cell counts. *Bottom Left*, quantitation of GFP^+^ frequency in host cardiac CD169^+^Tim4^+^ macrophages and total CD169^+^Tim4^+^ cells as a percentage of all autofluorescent(Auto)^+^ macrophages in vehicle and DT treated host MI mice; n=3-4/group, statistics: unpaired *t* test. ***C.*** *Left*, Parabiosis schema joining CD169^DTR^ host mice and either spleen-intact or Spx MaFIA donor parabionts, with host mice given DT at the time of MI to deplete CD169^+^ macrophages. *Right*, Example FACS dot plots and quantitation of donor CD45^+^Auto^+^CD64^+^MHCII^+^GFP^+^CD169^+^Tim4^+^ macrophages in the host MI heart 48 h post MI; n=3-4/group, statistics: unpaired *t* test.

For more comprehensive validation, we examined host CD169^DTR^ mice expressing human diphtheria toxin (DT) receptor (DTR) in the sialoadhesin gene allowing selective depletion of CD169^+^ macrophages^13^ and parabiont MaFIA mice expressing GFP in all mononuclear phagocytes, and used gating strategy 1 in *<u>Figure S3B</u>* to identify cardiac CD169^+^Tim4^+^ macrophages. In the first set of experiments, host CD169^DTR^ mice received either vehicle or DT (10 μg/kg i.p.) at the time of MI, and host cardiac leukocytes were examined 2 d later. As shown in *<u>Figure 3B</u>*, as compared to vehicle, DT-treated host mice exhibited robust increases in the frequency of parabiont-derived cardiac GFP^+^ leukocytes and GFP^+^CD169^+^Tim4^+^ macrophages, despite a reduction in total CD169^+^Tim4^+^ cells (expressed as percent of total cardiac macrophages). These data establish increased sourcing of cardiac CD169^+^ macrophages from the parabiont circulation after CD169^+^ cell ablation in the host. In the second set of studies, host CD169^DTR^ mice receiving DT at the time of MI were paired with either intact or Spx MaFIA parabionts, and host GFP^+^CD169^+^ cardiac macrophages were measured after 2 d. As compared to CD169^DTR^ mice paired with intact MaFIA parabionts, the hearts of host CD169^DTR^ mice paired with Spx MaFIA mice exhibited profoundly reduced GFP^+^CD169^+^ macrophages (*<u>Figure 3C</u>*). These data establish splenic dependence of augmented cardiac CD169^+^ macrophages from the parabiont circulation upon host CD169^+^ cell ablation.

We next evaluated the impact of MI, and the spleen, on cardiac CD169^+^Tim4^+^ macrophage LYVE1 subsets, using gating strategy 1 in *<u>Figure S3B</u>*. In naïve hearts, LYVE1^hi^ resident cells comprised the majority of CD169^+^Tim4^+^ macrophages (∼60%) versus LYVE1^low^ (*<u>Figures 4A and 1E</u>*). One day post-MI, this ratio reversed, with a marked decrease (∼1.8-fold) in LYVE1^hi^ frequency (consistent with ischemic loss^17, 45^) and ∼3.5-fold increase in LYVE1^low^ macrophages (*<u>Figure 4A</u>*). Bulk RNAseq of sorted LYVE1^hi^ and LYVE1^low^ cardiac CD169^+^Tim4^+^ macrophages 1 d post-MI revealed 462 DEGs (q<0.05) and subset segregation via hierarchical clustering (*<u>Figure 4B</u>*). GO pathway analysis revealed higher expression of metabolism and oxidative phosphorylation genes and lower expression of immune response genes in the LYVE1^low^ subpopulation in the acutely infarcted heart (*<u>Figure S7A</u>*), suggesting an immunomodulatory and pro-resolving phenotype. Moreover, gene expression analysis of the LYVE1^low^ subset (versus LYVE1^hi^ cells) revealed upregulation of several genes related to tissue injury (*Egr1*), wound healing, cell migration, and matrix responses (*Thbs1, Nrp1, Mmp9, Mmp12*), and immunomodulation (*Socs3, Sem4a*) consistent with a reparative and healing role for CD169^+^Tim4^+^ LYVE1^low^ macrophages (*<u>Figure S7B</u>*). These differences were much more pronounced post-MI as compared with naïve conditions. Interestingly, PCA of RNAseq data from LYVE1^hi^ and LYVE1^low^ CD169^+^ cardiac and splenic macrophages from naïve and post-MI mice revealed tight clustering of splenic cells regardless of underlying condition, but distinct clustering of cardiac macrophages linked to underlying injury, suggesting an overriding influence of tissue microenvironment on CD169^+^ macrophage phenotype (*<u>Figure 4C</u>)*. Importantly, Spx mice failed to expand CD169^+^Tim4^+^LYVE1^low^ cardiac macrophages after acute MI, exhibiting profoundly (∼11-fold) diminished levels of these cells compared to WT MI mice (*<u>Figures 4D-E</u>*). Collectively, these data establish that splenic CD169^+^Tim4^+^ MMMs with augmented phagocytic capacity and pro-resolving phenotype acutely traffic to the heart post-MI in response to chemotactic signals, and, analogous to steady state conditions, are a primary source of CD169^+^Tim4^+^LyVE1^low^ macrophages in the infarcted heart.

**Figure 4.**
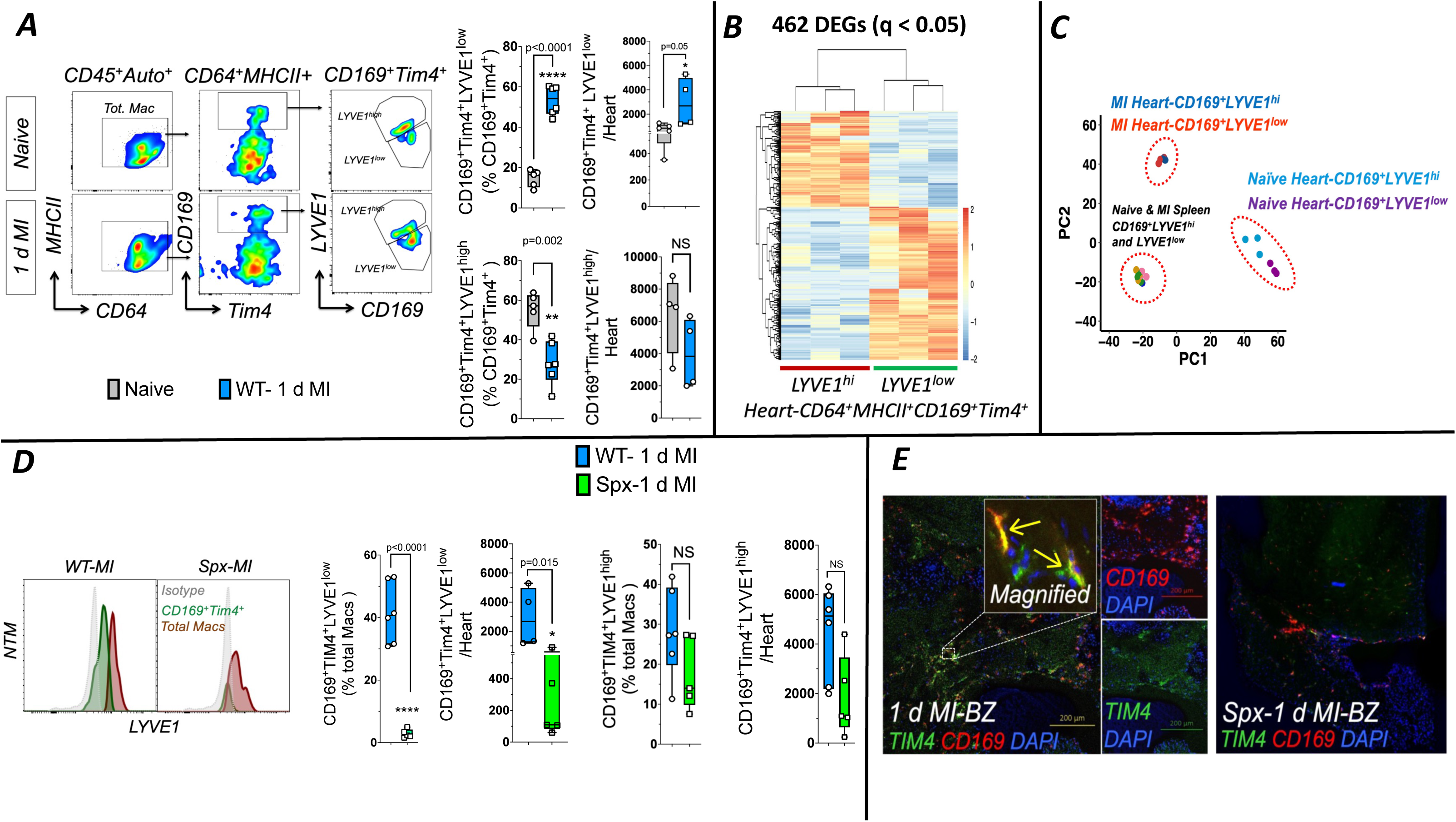
Spleen-derived CD169^+^Tim4^+^LYVE1^low^ macrophages expand in the acutely infarcted heart. ***A***. *Left,* FACS pseudocolor plots of cardiac CD169^+^Tim4^+^ macrophages and separation based on LYVE1 surface expression in naïve and 1 d post-MI mice. *Right*, quantitation of LYVE1^hi^ and LYVE1^low^ CD169^+^Tim4^+^ macrophages in hearts from naïve and 1 d post-MI mice. n=4-6/group; statistics: unpaired *t* test. ***B***. Heat map of 462 significant (p adjusted<0.05) DEGs by RNAseq analysis in sorted LYVE1^hi^ and LYVE1^low^ CD169^+^Tim4^+^ cardiac macrophages 1 d post-MI. ***C.*** PCA plots using the top 500 DEGs after rlog transformation of RNAseq data from these macrophages sorted from the indicated sites in naïve and 1 d post-MI mice. ***D.*** *Left*, Flow histograms depicting LYVE1 surface expression on cardiac CD169^+^Tim4^+^ macrophages (green) and total Autofluorescence^+^CD64^+^MHCII^+^ macrophages (brown) in WT and Spx mice, 1 d post-MI. *Right*, FACS quantitation of LYVE1^hi^ and LYVE1^low^ CD169^+^Tim4^+^ macrophages in the hearts of WT and Spx mice, 1 d post-MI; n=5-6/group. Statistics: unpaired *t* test. ***E.*** Representative confocal micrograph of border zone (BZ) myocardium immunostained for CD169 (red) and Tim4 (green) 1 d post-MI in WT and Spx mice; nuclear staining with DAPI (blue). Scale bar, 200 μm. Inset shows magnified images of CD169 and Tim4 staining. Yellow arrows indicate double positive cells; scale bar 10 μm.

### CD169^+^ macrophages and the spleen promote apoptotic neutrophil clearance, suppress neutrophil activation, and resolve inflammation post-MI

Coronary ligation was performed in WT mice, Spx mice, and CD169^DTR^ mice (given DT 1 h after ligation), with evaluation 24 h later. Relative to WT mice, both Spx mice and CD169^DTR^/DT mice exhibited marked reductions in blood Ly6C^low^CD169^+^Tim4^+^ cells (and total Ly6C^low^ monocytes) after acute MI (*<u>Figure 5A</u>*), without changes in Ly6C^hi^ monocytes (*<u>Figure S8A</u>*). Similarly, levels of Ly6C^low^CD169^+^Tim4^+^ macrophages and total Ly6C^low^ cells in the heart 24 h post-MI were profoundly reduced in both Spx mice and CD169^DTR^/DT mice (*<u>Figure 5B</u>*), again without differences in Ly6C^hi^ cells (*<u>Figure S8B</u>*). In contrast, as compared with WT, both Spx mice and CD169^DTR^/DT mice exhibited significantly elevated levels of blood neutrophils (and activated ICAM^+^ neutrophils) 24 h after MI (*<u>Figure 5C</u>*), indicative of heightened acute inflammation.

**Figure 5.**
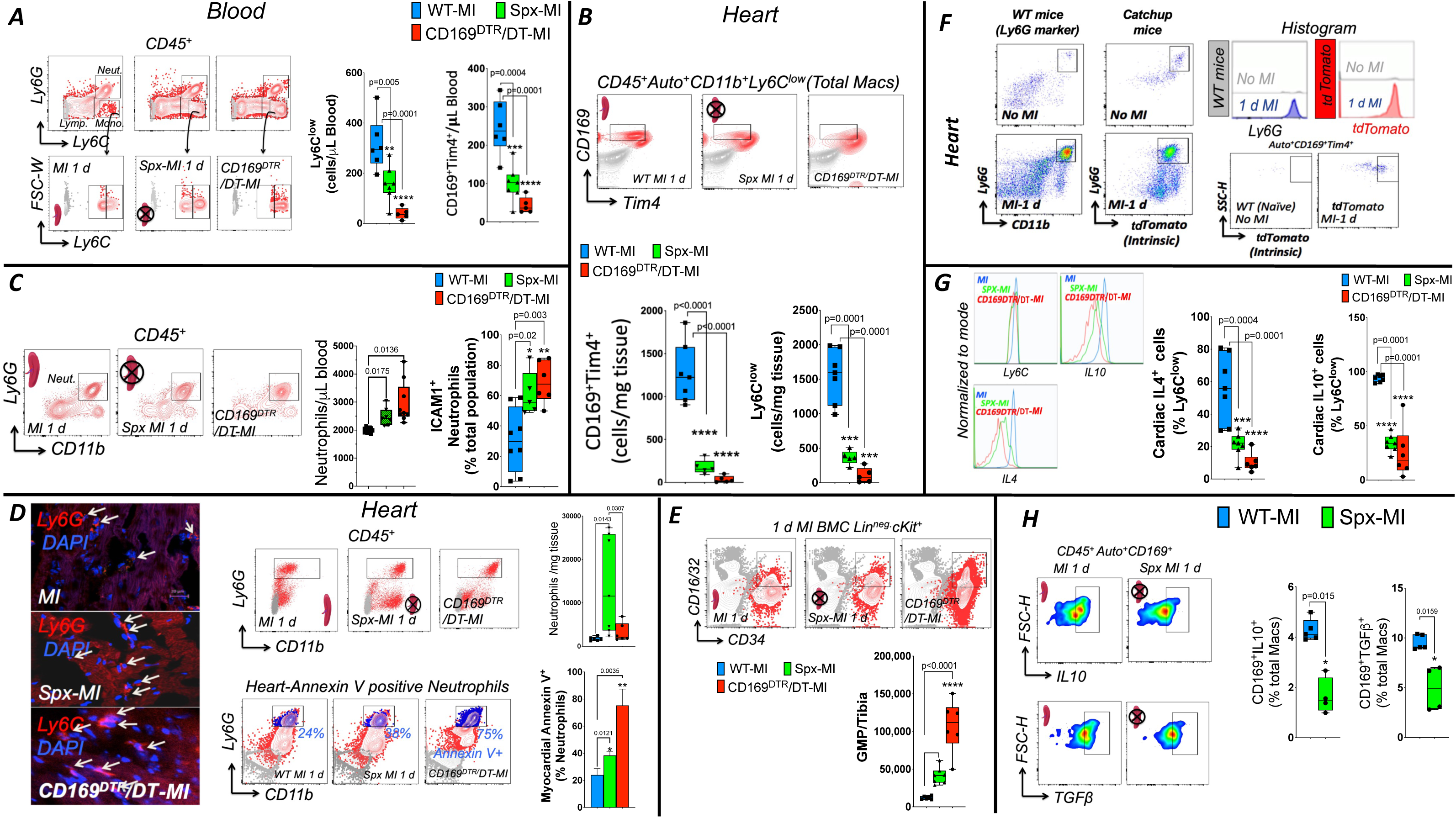
CD169^+^Tim4^+^ macrophages clear apoptotic neutrophils, suppress neutrophil activation, and resolve inflammation after MI. FACS plots and quantitation of Ly6C^low^CD169^+^Tim4^+^ macrophages and total Ly6C^low^ cells in blood (***A***) and in heart (***B***) 1 d post-MI in wild-type (WT) and splenectomized (Spx) WT mice, and CD169^DTR^ mice given diphtheria toxin (CD169^DTR^/DT) at the time of MI; n=5-7/group, statistics: one-way ANOVA, Bonferroni post-test. Isotype antibody is shown in gray. ***C*.** FACS plots and group data for blood CD45^+^CD11b^+^ Ly6G^+^ neutrophils and ICAM-1/CD54^+^ neutrophils 1 d post MI in WT, Spx, and CD169^DTR^/DT mice; n=6-8/group; statistics: one-way ANOVA, Bonferroni post-test. ***D*.** *Left,* Representative confocal images of immunofluorescent Ly6G staining in WT, Spx, and CD169^DTR^/DT hearts 1 d post-MI demonstrating Ly6G^+^ neutrophil (red) infiltration (arrows); nuclear staining with DAPI (blue). Scale bar 20 μm. *Right*, FACS plots and corresponding quantitation of cardiac CD45^+^CD11b^+^Ly6G^+^ neutrophils (red) and annexin V^+^ apoptotic neutrophils (blue) in the same groups 1 d post-MI; n=3-4/group, statistics: one-way ANOVA, Dunnett’sT3 post-test. ***E***. FACS density plots for Lin^−^c-kit^+^CD34^+^CD16/32^+^ granulocyte monocyte precursors (GMPs) in bone marrow from WT, Spx, and CD169^DTR^/DT mice 1 d post MI, together with quantitation. Flow gates were based on isotype antibody control. n=5-7/group; statistics: one-way ANOVA, Tukey’s post-test. ***F***. *Left*, Representative FACS dot plots identifying cardiac neutrophils as CD11b^+^Ly6G^+^ cells in WT mice and as Ly6G^+^tdTomato^+^ cells in Catchup mice at baseline and 1 d post-MI. *Right Top*, Representative histograms of Ly6G and tdTomato fluorescence intensity in heart mononuclear cells from the same groups. *Right Bottom*, FACS dot plots gated on cardiac CD169^+^Tim4^+^ macrophages illustrating tdTomato expression in Catchup mice 1 d post-MI. Auto, autofluorescence. ***G***. Representative FACS histograms of intracellular IL4 and IL10 staining in cardiac Ly6C^low^ cells 1 d post MI in WT, Spx and CD169^DTR^/DT mice, together with cell quantitation of the Ly6C^low^ subsets. N=6-7/group; statistics: one-way ANOVA, Bonferroni post-test. ***H***. Representative FACS pseudocolor plots for intracellular TGFβ and IL10 staining in cardiac CD169^+^ macrophages 1 d post MI in WT and Spx mice, with accompanying quantitation; n=4-5/group, statistics: unpaired *t* test.

Macrophage ingestion of apoptotic cells suppresses the production of pro-inflammatory mediators and promotes secretion of pro-resolving cytokines including interleukin(IL)-10 and transforming growth factor(TGF)-β.^50, 51^ During acute MI, apoptotic neutrophils undergo efferocytosis by macrophages.^1, 7^ As shown in <u>*Figure 5D and Figure S8C*</u>, as compared with WT, both Spx and CD169^DTR^/DT mice exhibited augmented cardiac neutrophil infiltration and increased annexin V^+^ apoptotic neutrophils after acute MI, as assessed by immunostaining and FACS. Neutrophilia in Spx and CD169^DTR^/DT mice was accompanied by increased Lin^−^ cKit^+^CD34^+^CD16/32^+^ GMPs in the bone marrow, suggesting augmented granulopoiesis upon loss of splenic MMMs (*<u>Figure 5E</u>*). To establish a role for CD169^+^Tim4^+^ macrophages in neutrophil efferocytosis, experiments were performed in Catchup mice,^24^ which contain a knock-in allele expressing Cre recombinase and the fluorescent protein tdTomato in the first exon of Ly6G. As depicted in *<u>Figure 5F</u>*, tdTomato positive neutrophils were identified in the heart at 24 h post-MI in these mice; ∼50% of CD169^+^Tim4^+^ macrophages in the acutely infarcted heart also exhibited tdTomato fluorescence, indicating the presence of ingested apoptotic neutrophils.

Reduced neutrophil efferocytosis in the absence of splenic CD169^+^Tim4^+^ macrophages post-MI would delay tissue macrophage polarization towards a reparative phenotype. Indeed, flow cytometric evaluation of cardiac Ly6C^low^ cells 24 h post-MI indicated profoundly reduced anti-inflammatory IL-10-and IL-4-expression in Spx and CD169^DTR^/DT mice as compared with WT mice (*<u>Figure 5G</u>*). Similarly, CD169^+^ macrophages in the acutely infarcted heart exhibited ∼2-fold lower IL-10^+^ and TGF-β^+^ cells in Spx MI mice as compared to WT MI (*<u>Figure 5H</u>*), and F4/80 and IL-10 immunostaining of the infarct border zone revealed significantly fewer dual F4/80^+^IL-10^+^ (yellow) cells in Spx MI mice (*<u>Figure S8D</u>*). Taken together, these studies demonstrate that CD169^+^Tim4^+^ macrophages, including those of splenic origin, temper the neutrophil inflammatory response and promote efferocytosis and the transition to a pro-resolving milieu in the heart after acute MI.

### Splenic CD169^+^Tim4^+^ macrophages promote post-MI healing and reduce adverse LV remodeling

WT mice, Spx mice, and CD169^DTR^/DT mice underwent non-reperfused MI or sham operation and followed for 10 d. Compared to respective sham mice, all MI mice exhibited increased mortality over 10 d (*<u>Figure 6A</u>*). However, post-MI mortality was dramatically, and comparably, higher in both Spx mice and CD169^DTR^/DT mice versus WT mice. Necropsy revealed increased mortality due to more frequent cardiac rupture (*<u>Figure 6B</u>*), consistent with impaired post-MI wound healing in both Spx and CD169^DTR^/DT mice.

**Figure 6.**
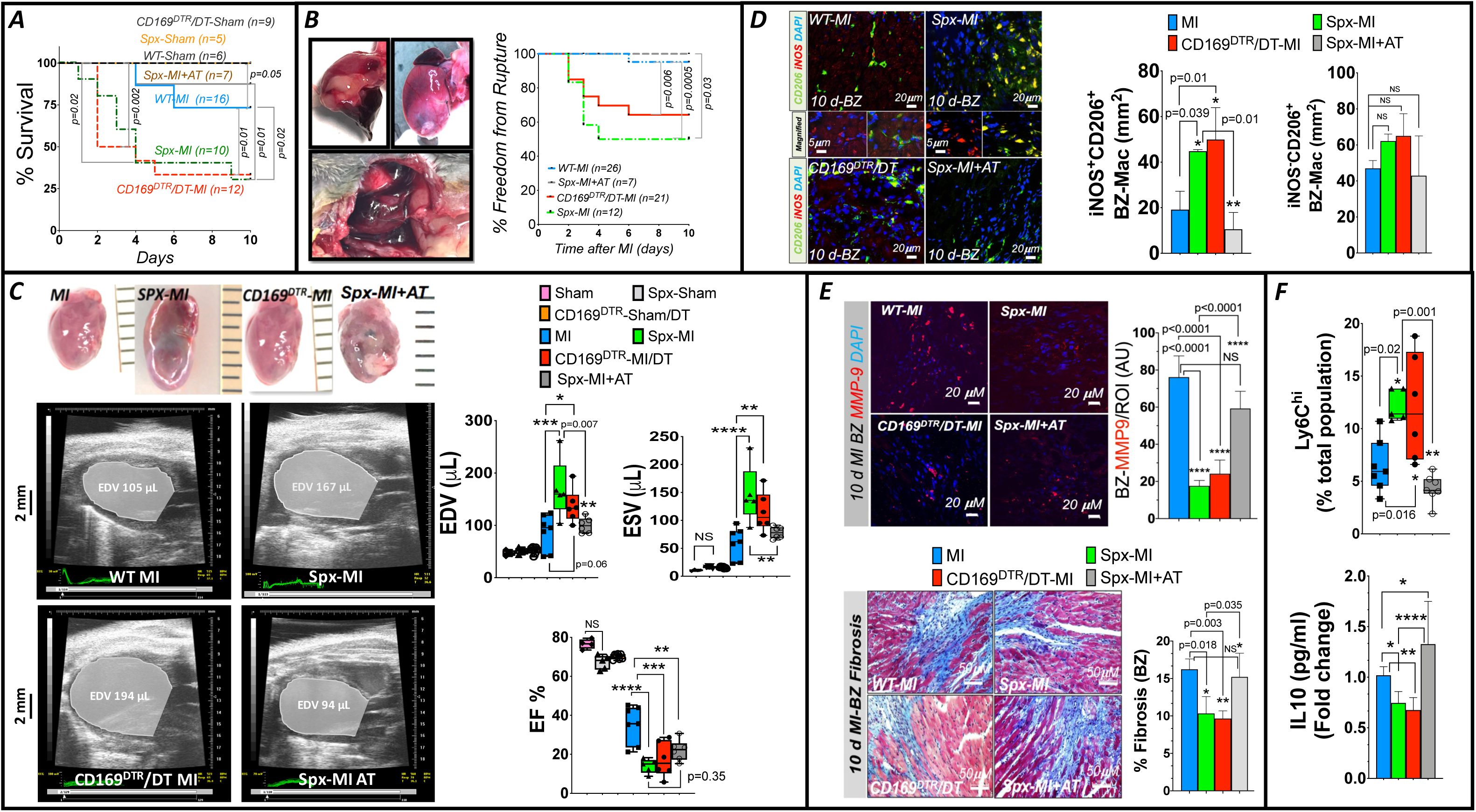
Splenic CD169^+^Tim4^+^ marginal metallophilic macrophages promote healing and reduce adverse LV remodeling after MI. ***A*.** Kaplan-Meier survival curves over 10 d following MI or sham operation in WT, Spx, and CD169^DTR^/DT mice, and after MI in CD45.2 Spx mice with adoptive transfer of naïve splenic CD169^+^Tim4^+^ cells from syngeneic CD45.1 WT mice 24 h post-MI (Spx-MI+AT). Statistical comparisons: log-rank test. ***B***. Gross images of post-MI cardiac rupture with hemothorax or hemopericardium, and Kaplan-Meier curves for freedom from rupture over 10 d post-MI in WT, Spx, CD169^DTR^/DT, and Spx-MI+AT mice. Statistical comparisons: log-rank test. ***C*.** *Left*, Representative post-mortem whole hearts and end-diastolic long-axis 2-dimensional echocardiograms from WT-MI, Spx-MI, CD169^DTR^/DT-MI and Spx-MI+AT mice (10 d post-MI). *Right*, Quantitation of LV ejection fraction (EF) and end-diastolic and end-systolic volume (EDV and ESV) at 10 d post-MI; n=4-6/group, statistics: one-way ANOVA, Bonferroni post-test. ***D*.** *Left*, Representative confocal images of immunofluorescent staining for CD206 (green) and iNOS (red) in the heart infarct border zone (BZ) from WT-MI, Spx-MI, CD169^DTR^/DT-MI, and Spx-MI+AT mice 10 d post-MI. DAPI (blue) nuclear staining. iNOS^+^CD206^+^ cells appear yellow. Higher magnification is shown in the middle panel. *Right*, quantitation of iNOS^+^CD206^+^ and iNOS^−^CD206^+^ cells/mm^2^ in the hearts. N=3/group, statistics: one-way ANOVA, Bonferroni post-test. NS, not significant. ***E*.** *Top Left*, Confocal images of MMP-9 immunostaining (red) in infarct BZ of hearts from WT-MI, Spx-MI, CD169^DTR^/DT-MI, and Spx-MI+AT mice 10 d post-MI. Nuclear staining with DAPI (blue). *Top Right*, Quantitative group data for total MMP-9 mean fluorescence intensity per region of interest (ROI). AU, arbitrary units. N=4/group, statistics: one-way ANOVA, Bonferroni post-test. *Bottom*, Representative Masson’s trichrome stains of infarcted hearts (10 d post-MI) from the same groups demonstrating MI border zone (BZ) fibrosis, together with BZ fibrosis quantitation. n=4/group; statistics: one-way ANOVA, Bonferroni post-test. ***F***. FACS quantitation of Ly6C^hi^ blood monocytes and serum IL-10 levels in WT-MI, Spx-MI, CD169^DTR^/DT-MI, and Spx-MI+AT mice at 10 d post-MI. N=4-7/group, statistics: one-way ANOVA, Bonferroni post-test.

To assess the sufficiency of splenic MMMs for cardiac repair, CD45.2 WT Spx mice underwent MI, but with intravenous adoptive transfer (AT) 24 h after MI of 1×10^6^ FACS-sorted CD169^+^Tim4^+^ cells (∼100 μL/mouse) from the spleens of donor CD45.1 WT mice (*Figure S9A*).

Post-AT (24 h), CD45.1^+^CD169^+^Tim4^+^ macrophages were readily identified in the heart and blood by immunostaining and flow cytometry (*<u>Figure S9B</u>*), indicating successful donor cell transfer, with recipient Spx MI mice augmenting circulating Ly6c^low^CD169^+^Tim4^+^ cells to levels comparable to non-splenectomized MI mice (*<u>Figure S9C</u>*). Notably, splenic CD169^+^Tim4^+^ cell reconstitution rescued the increased mortality and rupture rate in Spx MI mice (*<u>Figure 6A-B</u>*), reduced bone marrow neutrophil levels (*<u>Figure S9D</u>*), and markedly diminished the abundance (∼13-fold) of both total and annexin V^+^ neutrophils in the infarcted heart 24 h post-transfer (*<u>Figure S9E</u>*) indicating that splenic MMMs are both necessary and sufficient for tissue repair, temper acute neutrophil inflammation, and augment apoptotic neutrophil efferocytosis. Loss of these specialized macrophages contributes critically to deleterious post-MI healing in Spx mice.

Echocardiography at 10 d post-MI revealed significantly larger LV end-diastolic and end-systolic volumes (EDV and ESV) and lower LV ejection fraction (EF) in Spx and CD169^DTR^/DT MI mice versus WT MI mice (*<u>Figure 6C</u>*), indicative of exacerbated LV remodeling. Splenic MMM reconstitution in Spx MI mice improved LV remodeling, evidenced by significantly lower EDV and ESV (comparable to WT MI) versus Spx MI mice (*<u>Figure 6C</u>*). Reparative CD206^+^ macrophages are an important cardiac macrophage population 10 d post-MI.^1, 6, 52^ Although CD206^+^ cells were readily identifiable upon border zone immunostaining in all mouse MI groups (*<u>Figure 6D</u>*), both Spx and CD169^DTR^/DT MI mice exhibited a ∼2.5-fold increase in CD206^+^ macrophages co-expressing pro-inflammatory iNOS. Splenic MMM transfer in Spx mice markedly decreased (∼4-fold) iNOS^+^CD206^+^ cardiac macrophages post-MI to levels similar to WT mice. Abundance of cardiac iNOS^−^CD206^+^ cells was comparable across all MI groups (*<u>Figure 6D</u>*). Accompanying this persistent pro-inflammatory milieu, there was significantly less border zone matrix metalloproteinase-9 (MMP9) abundance and fibrosis 10 d post-MI in both Spx and CD169 ablated MI mice, with restoration of MMP-9 and border zone fibrosis upon splenic MMM transfer in Spx MI mice (*<u>Figure 6E</u>*). These tissue level changes were accompanied by parallel directional changes in pro-inflammatory Ly6C^hi^ monocytes and serum levels of anti-inflammatory IL-10 (*<u>Figure 6F</u>*). In a subgroup of WT, Spx, and CD169^DTR^/DT mice, long-term (8 w) post-MI remodeling was examined (*<u>Figure S10</u>*). As compared to WT MI, Spx and CD169^DTR^/DT MI mice exhibited: 1) aggravated LV remodeling and HF, with increased chamber volume, greater heart and lung weight, and lower EF; 2) thinner scars and reduced border zone fibrosis, and 3) chronic inflammation with persistently augmented iNOS^+^CD206^+^ cardiac macrophages.

Collectively, these findings indicate that splenic CD169^+^Tim4^+^ macrophages are indispensable for efferocytosis and inflammation resolution, matrix turnover, wound healing and scar formation, and effective remodeling of the post-MI heart. Deficiency of CD169^+^ macrophages early post-MI exacerbates LV rupture, adverse long-term LV remodeling, and late HF.

### Splenic marginal zone expansion ameliorates adverse LV remodeling and inflammation after MI

Liver X receptor-α (LXRα) is a nuclear receptor essential for the generation of splenic MZ macrophages. LXRα enhances macrophage phagocytosis and suppresses inflammatory pathways after efferocytosis.^53, 54^ In naïve mice, the selective LXRα agonist T0901317^55^ (40 mg/kg i.p.) induced a ∼9-fold increase in blood CD169^+^Tim4^+^ macrophages and a ∼1.5-fold expansion of splenic CD169^+^Tim4^+^ MMMs 24 h later (*<u>Figure S11</u>*). To evaluate the effects of splenic MZ expansion during MI, we administered T0901317 40 mg/kg i.p daily from 1 d prior to 5 d after MI in WT and Spx mice (*<u>Figure 7A</u>*). T0901317 increased CD169^+^Tim4^+^ macrophages in the heart 24 h post-MI and blood CD169^+^Tim4^+^ macrophages10 d post-MI in WT MI mice, but not in Spx MI mice (*<u>Figure 7B)</u>*. Importantly, T0901317-treated WT MI mice exhibited significantly improved 10 d post-MI mortality as compared with untreated MI mice (*<u>Figure 7C</u>*) but did not improve post-MI mortality in Spx mice. The mortality benefit resulted primarily from reduced ventricular rupture, consistent with improved wound healing.

**Figure 7.**
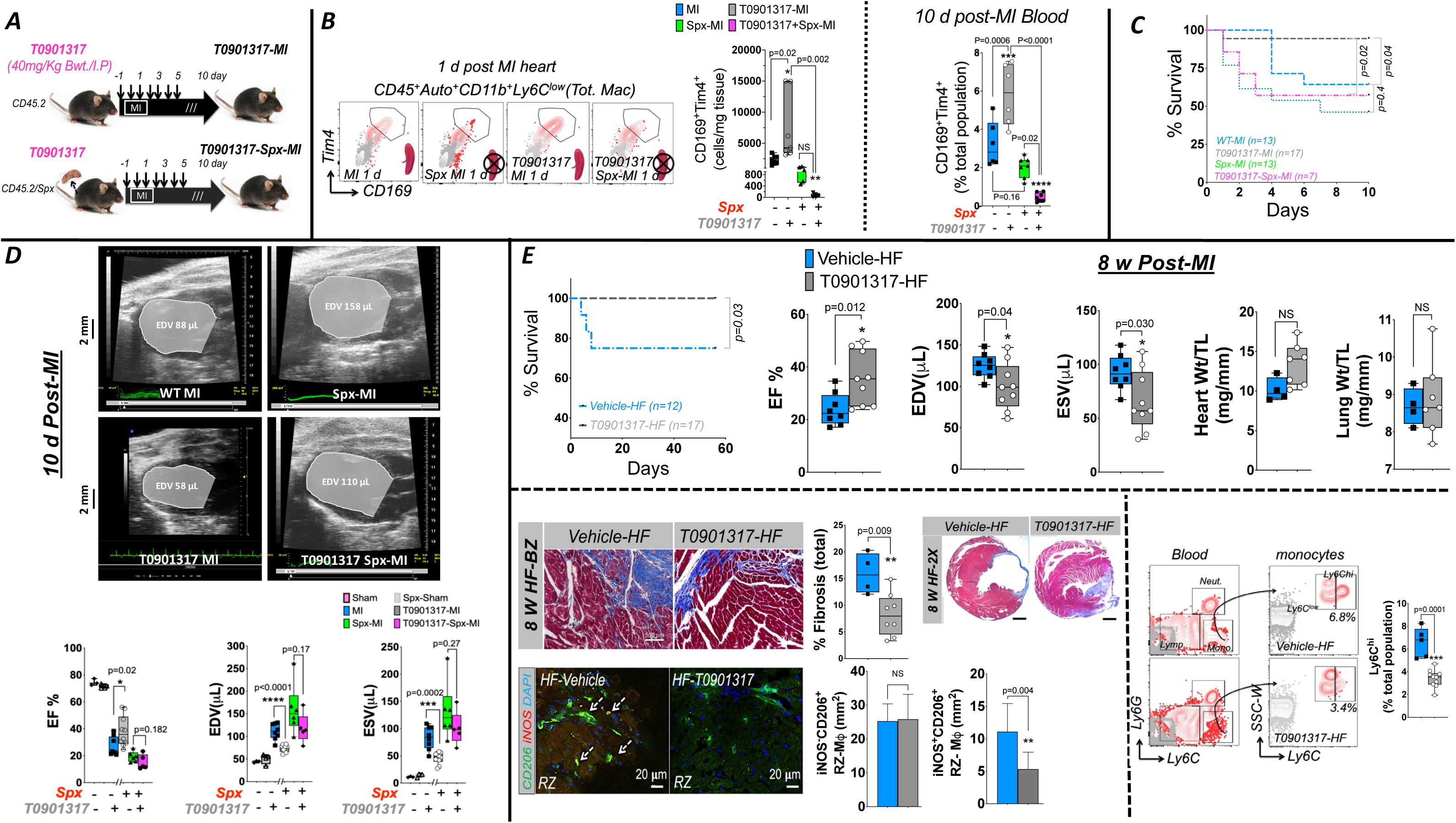
Splenic marginal zone expansion alleviates LV remodeling and inflammation after MI. ***A*.** Protocol for LXRα agonist T0901317 treatment (40 mg/kg i.p.) from 1 d prior to 5 d post-MI, with 10 d post-MI follow-up, in WT and Spx mice. ***B*.** FACS contour plots and quantitation of cardiac CD169^+^Tim4^+^ macrophages 1 d post-MI and blood CD169^+^Tim4^+^ macrophages 10 d post-MI in untreated and T0901317-treated WT and Spx mice. N=5-6/group, statistics: one-way ANOVA, Bonferroni post-test. ***C*.** Kaplan-Meier survival curves post-MI in untreated and T0901317-treated WT and Spx mice. Statistical comparisons by log-rank test, group sizes as indicated. ***D*.** Representative end-diastolic long-axis 2-dimensional echocardiograms and group data for LV ejection fraction (EF) and end-diastolic and end-systolic volume (EDV and ESV) in the same mouse groups at 10 d post-MI; n=5-10/group, statistics: one-way ANOVA, Bonferroni post-test. ***E*.** *Top*, Kaplan-Meier survival curves over 8 w post-MI in WT mice treated with either vehicle or T0901317 from 1 d before MI to 5 d post-MI (statistical comparison by log-rank test, n=12-17/group as indicated) and group data for LVEF, LVEDV, LVESV, and normalized heart and lung weight at 8 w post-MI; n=8-9/group for echocardiography, n=4-7/group for gravimetry. Statistical comparisons: unpaired *t* test. HF, heart failure; TL, tibia length; NS, not significant. *Bottom Left*, Representative Masson’s trichrome staining of LV short-axis sections (2x magnification) and infarct border zone (BZ, scale bar 500 μm), along with quantitation of cardiac fibrosis (BZ and remote zone [RZ], blue staining) in vehicle- and T0901317-treated WT HF mice. Also shown are confocal images of immunofluorescent staining for CD206 (green) and iNOS (red) in the RZ of hearts from vehicle-and T0901317-treated HF mice (8 w post-MI) and quantitation of iNOS^+^CD206^+^ and iNOS^−^ CD206^+^ macrophages (Mφ) per mm^2^. Double positive (CD206^+^iNOS^+^) cells appear yellow (arrows). DAPI (blue) was used for nuclear staining. N=4-8/group, statistics: unpaired *t* test. *Bottom Right*, Representative FACS contour plots to identify Ly6C^hi^ monocytes in vehicle- and T0901317-treated WT HF mice (8 w post-MI), and corresponding quantitation. N=5-10/group, statistics: unpaired *t* test.

At 10 d post-MI, T0901317-treated WT MI mice exhibited significantly smaller EDV and ESV and higher EF than untreated WT MI mice (*<u>Figure 7D</u>*). In contrast, there was no effect of T0901317 on LV volumes and EF in Spx MI mice. Evaluation of a separate group of WT MI mice 8 w post-MI revealed that T0901317 given early after MI significantly improved (versus vehicle) long-term survival, LVEDV, LVESV, and LVEF (*<u>Figure 7E</u>*). There was less total cardiac interstitial fibrosis (border and remote zone), and fewer pro-inflammatory iNOS^+^CD206^+^ macrophages and circulating Ly6C^hi^ monocytes with T0901317 treatment, but comparable levels of iNOS^−^CD206^+^ cardiac macrophages as compared to vehicle-treated MI mice (*<u>Figure 7E</u>*). Taken together, these results establish splenic dependence of the remodeling benefits of the LXRα agonist T0901317 and that marginal zone macrophage expansion is cardioprotective post-MI, suppressing cardiac inflammation and improving short- and long-term cardiac remodeling.

### Blood CD14^+^HLA-DR^+^CD169^+^ monocytes expand after ST-segment elevation MI (STEMI) in humans

After informed consent, blood was collected from patients presenting with acute STEMI prior to urgent coronary reperfusion, and from control subjects with coronary artery disease prior to undergoing elective PCI. The two groups were matched for age, sex, and race, and had comparable co-morbidity burden, as depicted in *<u>Table S1</u>*. Using the gating strategy in *<u>Figure 8A</u>,* we identified CD169^+^Tim4^+^ macrophages within the circulating CD14^+^HLA-DR^+^CD64^+^ cell population. As circulating CD169^+^ macrophages have not, to our knowledge, previously been described in humans, we performed further validation by sorting CD14^+^HLA-DR^+^CD169^+^ cells and visualizing them with imaging flow cytometry (*<u>Figure 8B</u>*). Robust CD169 expression was readily observed in sorted cells. Moreover, circulating CD169^+^ macrophages from STEMI subjects exhibited a narrower size distribution (∼9 μm) as compared with control PCI (8-11 μm) (*<u>Figure 8B)</u>*, suggesting an altered activation state. Quantitation by flow cytometry revealed a robust increase in the frequency of CD169^+^ and CD169^+^Tim4^+^ circulating macrophages in STEMI subjects versus PCI controls (*<u>Figure 8C</u>*). Taken together, these data suggest mobilization of activated CD169^+^ macrophages after acute STEMI in humans.

**Figure 8.**
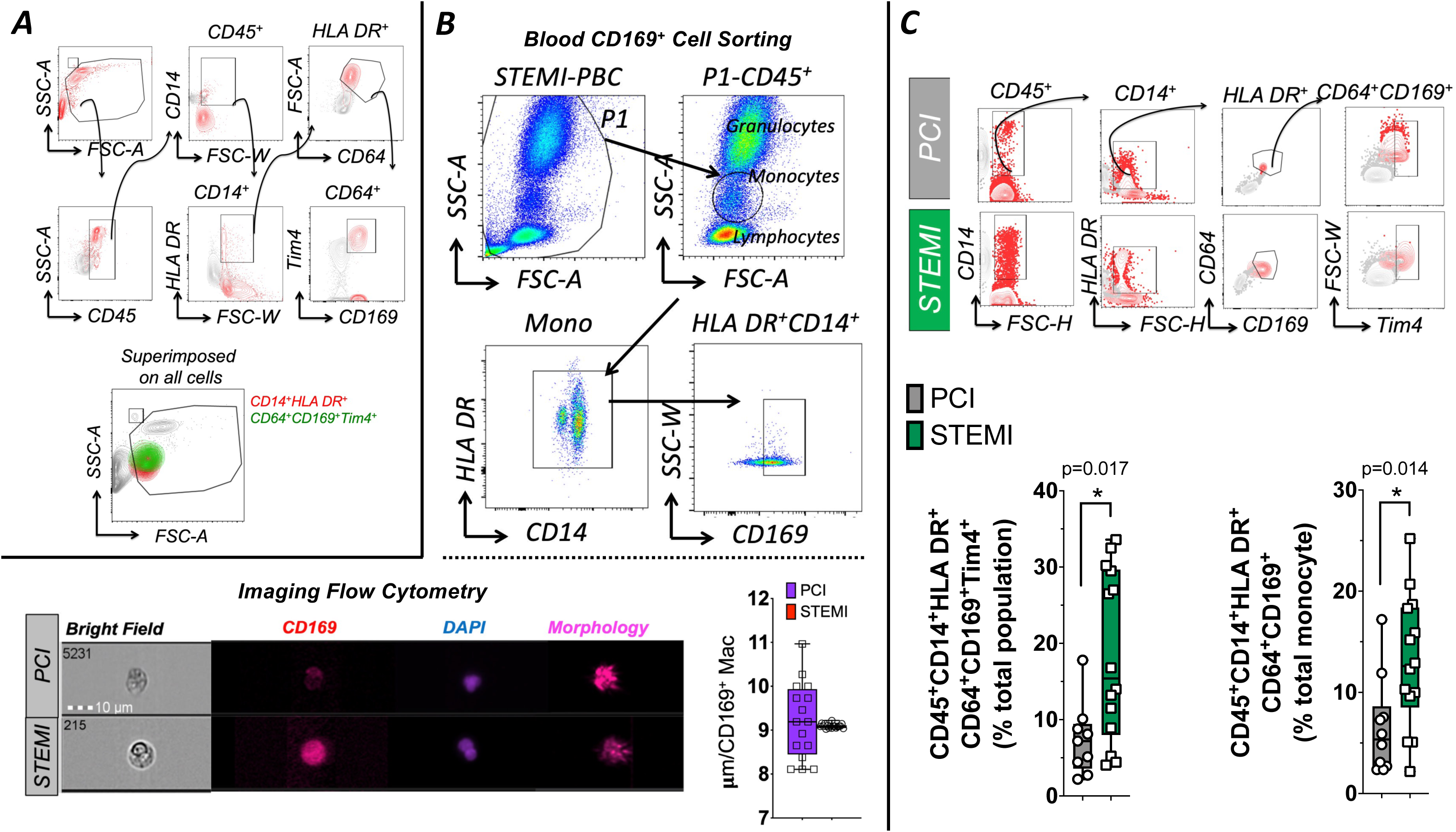
CD169^+^Tim4^+^ macrophage expansion after ST-segment elevation MI (STEMI) in humans. ***A*.** FACS plots and characterization of human CD45^+^CD14^+^HLA- DR^+^CD64^+^CD169^+^Tim4^+^ circulating macrophages from a subject with acute STEMI. The accompanying overlaid contour plot illustrates CD14^+^HLA-DR^+^ (red) and CD64^+^CD169^+^Tim4^+^ (green) subsets superimposed on all CD45^+^ leukocytes in an SSC-A versus FSC-A gate. ***B*.** *Top*, FACS gating strategy for sorting CD45^+^CD14^+^HLA-DR^+^CD169^+^ cells from human peripheral blood for further characterization using ImageStream analysis. *Bottom*, ImageStream visualization of FACS-sorted CD45^+^CD169^+^ blood cells from STEMI patients and control subjects undergoing elective percutaneous coronary intervention (PCI), and group data for size distribution of CD169^+^ cells; scale bar 10 μm. ***C*.** FACS contour plots and quantitation of circulating CD45^+^CD14^+^HLA-DR^+^CD64^+^CD169^+^Tim4^+^ macrophages in STEMI and control PCI subjects. n=11-14/group, statistics: unpaired *t* test.

## DISCUSSION

We have established a novel role for splenic CD169^+^Tim4^+^ metallophilic marginal macrophages (MMMs), and the cardiosplenic axis, in the inflammatory, healing, and remodeling responses after MI. There are several key findings. First, splenic CD169^+^Tim4^+^ MMMs circulate in blood as Ly6C^low^ cells and help maintain CD169^+^Tim4^+^CCR2^−^LYVE1^low^ macrophages in the naïve heart. Second, after acute MI, splenic CD169^+^Tim4^+^ MMMs augment phagocytosis and mobilize to the heart in response to chemotactic signals, resulting in marked expansion of CD169^+^Tim4^+^LyVE1^low^ macrophages with an immunomodulatory and pro-resolving gene signature. Third, splenic MMMs play a critical role in efferocytosis post-MI, particularly in clearance of apoptotic neutrophils, and are obligatory for inducing a reparative macrophage phenotype. Fourth, splenic CD169^+^Tim4^+^ macrophages are both necessary and sufficient for proper post-MI wound healing, and limit long-term pathological remodeling post-MI. Fifth, LXRα agonist-induced expansion of the splenic marginal zone and MMMs alleviates adverse post-MI LV remodeling and inflammation, suggesting a possible translational approach for splenic modulation. Lastly, analogous to mice, humans also exhibit expansion of circulating CD14^+^HLA-DR^+^CD64^+^CD169^+^Tim4^+^ macrophages after acute STEMI. Collectively, these data establish that splenic CD169^+^Tim4^+^ marginal metallophilic macrophages migrate to the heart and are required for the pro-resolving and reparative response after MI. Hence, these macrophages limit long-term pathological LV remodeling, and can potentially be manipulated for therapeutic benefit.

Interfacing the white and red pulp, the splenic marginal zone is an important area for the transit of blood leukocytes.^10^ It contains specialized CD169^+^ MMMs that clear cell debris and apoptotic cells through specialized phagocytic receptors such as Tim4,^16^ capture blood-borne antigens, and regulate subsequent adaptive immune responses.^56, 57^ CD169 is a lectin-like receptor whose expression is restricted to tissue-resident macrophages in various organs. The spatial localization of CD169^+^ macrophages in the splenic marginal zone, within hepatic sinusoids, and adjacent to the brain and kidney vasculature suggests they are frontline sentinels in phagocytosing and removing blood borne particulate antigens.^58–60^ Splenic marginal zone macrophages are required for the establishment of tolerance to apoptotic cell antigens,^32^ an essential step to reduce immunogenicity of autoantigens, as well for the amplification of immune responses through interactions with dendritic cells. These are consistent with a unique macrophage population that can regulate both tolerogenic and pro-inflammatory responses.

CD169^+^ MMMs are classically considered tissue resident at steady-state, dependent on CCL19 and CCL21 for splenic localization.^10, 57^ However, our data indicate that splenic CD169^+^Tim4^+^ macrophages (expressing MHCII and the core macrophage marker CD64, confirmed with lineage traced mice) also circulate in blood primarily as Ly6C^low^ cells, and populate the heart as CD169^+^Tim4^+^LyVE1^low^ cells. The circulating cells exhibited projections reminiscent of splenic MMM processes that penetrate the white pulp.^57^ Moreover, splenectomy induced an ∼80% loss of cardiac CD169^+^Tim4^+^ macrophages, consistent with an active spleno-cardiac MMM circulation. CD169^+^Tim4^+^ macrophages are particularly important as an immunoregulatory and hypo-stimulatory tissue-resident macrophage subset.^20^ Notably, while both splenectomy and CD169 ablation significantly reduced total Ly6C^low^ monocytes, neither impacted levels of pro-inflammatory Ly6C^hi^ monocytes.

Given a steady-state spleno-cardiac circulation of CD169^+^Tim4^+^ macrophages, the recognized rapid turnover of splenic MMMs during inflammatory states,^57, 61^ and their importance in immunoregulation,^20^ we explored their role in acute MI. It is established that splenic subcapsular red pulp reservoir Ly6C^hi^ monocytes infiltrate the heart and contribute to both inflammation and repair after acute MI.^2, 3, 5, 6, 9, 57, 61^ Here we show, to our knowledge for the first time, that Ly6C^low^CD169^+^ Tim4^+^ splenic MMMs also robustly mobilize to the heart after acute MI, as Spx dramatically reduced the cardiac recruitment of CD169^+^Tim4^+^ LYVE1^low^ macrophages, and also resulted in augmented donor splenic MMM recruitment in parabiotic models. CD169^+^Tim4^+^ MMMs exhibited increased surface expression of CCR3 and CCR4 in conjunction with upregulation of several corresponding chemokine ligands in the heart. Interestingly, spleen-derived MMMs augmented phagocytic capacity after MI, suggesting that efferocytosis is a key role for these cells.

CD169^+^Tim4^+^ MMMs were pro-resolving, anti-inflammatory, and reparative in the infarcted heart. An important mechanism underlying these effects may be their impact on neutrophil clearance and production. Tim4 binds phosphatidylserine residues expressed on the surface of apoptotic cells, allowing for proper engulfment and prevention of ambient release of toxic materials from dead cells.^16^ After acute MI, there is initial intense neutrophil infiltration for removal of tissue debris.^1, 6^ Neutrophils have short life spans and undergo apoptosis,^62^ necessitating efferocytosis of a large number of apoptotic neutrophils in the acutely infarcted heart. Our studies with Catchup mice established that CD169^+^Tim4^+^ macrophages take up apoptotic neutrophils, while MMM loss upon either splenectomy or CD169 ablation significantly increased the number of apoptotic neutrophils in the infarcted heart. Apoptotic neutrophil levels normalized upon adoptive transfer of splenic CD169^+^Tim4^+^ MMMs in Spx MI mice, establishing these cells as critical mediators of neutrophil efferocytosis after MI.

Apoptotic neutrophil disposal is an important step in inflammation resolution, as it triggers anti-inflammatory and immunosuppressive signals in engulfing phagocytes,^63^ including an IL-12^low^IL10^high^ regulatory phenotype^64^ that promotes tissue repair. In our studies, mice with MMM deficiency after splenectomy or CD169 ablation exhibited significantly fewer cardiac Ly6C^low^ cells expressing anti-inflammatory IL-10 or IL-4 24 h post-MI, more pronounced pro-inflammatory Ly6C^hi^ monocytosis and lower anti-inflammatory serum IL-10 levels at 10 d post-MI, and persistence of pro-inflammatory iNOS^+^CD206^+^ macrophages at later stages (10 d and 8w post-MI) of remodeling. Pro-inflammatory macrophage abundance, Ly6C^hi^ monocytosis and serum IL-10 levels were all normalized upon adoptive transfer of splenic CD169^+^Tim4^+^ MMMs in Spx MI mice. Hence, while the spleen is a complex organ whose physiological impact on the heart likely relates to multiple splenocyte cell types, these studies establish that splenic MMMs are essential for the development of a pro-resolving program in cardiac macrophages after MI.

Loss of splenic Ly6C^low^CD169^+^ Tim4^+^ cells also augmented neutrophil activity and accumulation in the heart thereby sustaining tissue inflammation. Both Spx and CD169 ablated mice exhibited increased total and ICAM1^+^ activated blood neutrophils, myocardial neutrophils, and bone marrow GMPs (suggesting greater granulopoiesis) acutely (24 h) after MI. Bone marrow and myocardial neutrophils were markedly suppressed upon splenic CD169^+^Tim4^+^ MMM adoptive transfer in Spx MI mice. These data are consistent with previous work showing that: 1) Ly6C^low^CD169^+^ macrophages limit kidney neutrophil accumulation and injury after ischemia-reperfusion by suppressing endothelial ICAM-1 expression on vascular endothelial cells;^65^ 2) mice deficient in marginal zone and bone marrow stromal cell macrophages due to myeloid C-FLIP deficiency exhibit excess G-CSF-driven granulopoiesis due to the failure of macrophages to efficiently clear apoptotic neutrophils;^66^ and 3) acute CD169^+^ macrophage depletion significantly expands an activated aged neutrophil subset.^67^ Hence, the anti-inflammatory and reparative effects of splenic Tim4^+^ MMMs appear tightly linked to neutrophil responses.

In concert with immunomodulatory effects, CD169^+^Tim4^+^ MMMs induced functional benefits in the infarcted heart, promoting cardiac wound healing and limiting early and late LV remodeling and dysfunction. Both Spx and CD169 ablated MI mice exhibited higher 10-day mortality after MI due to an increased rate of cardiac rupture, indicative of impaired wound healing. Lower survival was accompanied by lower MMP9 levels and reduced border zone fibrosis at 10 days post-MI, indicating diminished extracellular matrix turnover and healing scar formation, together with increased LV chamber dilatation and systolic dysfunction. Impressively, CD169^+^Tim4^+^ MMM adoptive transfer in Spx MI mice improved survival, prevented rupture, restored border zone MMP9 and fibrosis, and alleviated LV dilatation at 10 days post-MI. The long-term remodeling effects (8 weeks post-MI) of splenic MMM deficiency included thinner scars, and increased LV dilatation, systolic dysfunction, hypertrophy, and lung edema. These data indicate that splenic CD169^+^Tim4^+^ MMMs are obligatory for optimal post-MI wound healing to limit the late development of ischemic cardiomyopathy and HF.

It has been previously shown that splenectomized mice exhibit exaggerated early LV remodeling post-MI.^9^ Our results establish that this impaired healing response is in part dependent on splenic CD169^+^ MMMs. Our studies are also consistent with more recent work demonstrating that CD169^+^ macrophage depletion one week before reperfused MI worsened post-MI remodeling.^45^ In that study, CD169 was used as a marker of tissue resident macrophages to understand the impact of cardiac CCR2^−^ cells. Our results complement this prior work to indicate that detrimental effects of CD169^+^ macrophage ablation is also due to the loss of splenic MMMs, as non-splenic derived cardiac resident CD169^+^Tim4^+^ cells would not be impacted by splenectomy. Importantly, the pro-healing effects of the spleen on the heart during acute MI described herein contrasts with the detrimental cardiosplenic axis in chronic ischemic cardiomyopathy and heart failure that we have described previously.^26^ This suggests a pathological change in the splenic immune cell milieu in chronic HF that favors tissue injury rather than healing, and carries important translational implications for the wound healing following a first MI versus recurrent MI in the context of underlying ischemic cardiomyopathy and HF.

The above studies established the necessity of splenic MMMs for post-MI wound healing. We also wanted to establish whether expansion of splenic MMMs would improve endogenous post-MI repair. LXRs are nuclear receptors that are key transcriptional regulators of metabolic and immune responses in macrophages, including the phagocytic clearance of apoptotic cells and immune tolerance.^54, 68^ Among the two isoforms (LXRα and LXRβ), LXRα is specifically required for the development and generation of MMMs (and MZMs).^69^ LXRα deficient mice exhibit selective ablation of marginal zone macrophages, without impacting CD169^+^ macrophage populations in other tissues.^69^ Moreover, LXRα activation accelerates marginal zone macrophage development and expansion. Therefore, we used pharmacological LXRα agonism to expand splenic MMMs and their efferocytotic capacity, as a means to evaluate therapeutic benefit of MMM gain-of-function after MI. Since LXRs are expressed in several tissues, we isolated the splenic effects of LXRα agonism by comparing responses in WT and Spx mice. Notably, pharmacological LXRα augmentation with T0901317 robustly increased splenic and circulating CD169^+^Tim4^+^ cells at steady-state, and splenic CD169^+^Tim4^+^ MMM infiltration in the acutely infarcted heart. T0901317 improved 10-day post-MI survival and LV remodeling in WT mice only, and not in Spx mice, indicating the splenic dependence of these effects. Long-term effects included alleviation of late LV systolic dysfunction, total fibrosis, and reduction of pro-inflammatory monocytes and macrophages suggesting chronic suppression of both local and systemic inflammation.

LXRα agonism may be a potential translational approach to favorably modulate the spleen after MI. Our results extend previous studies demonstrating that both dual LXRα/β agonists^70, 71^ and LXRα-selective agonists^72^ protect against adverse LV remodeling from pressure-overload, whereas such remodeling is worsened in LXRα null mice.^71^ Bolstering the case for clinical relevance, analogous to our findings in mice with acute MI, humans with acute STEMI also exhibited marked expansion of circulating CD14^+^HLA-DR^+^CD64^+^CD169^+^Tim4^+^ macrophages, along with size distribution characteristics suggesting an altered activation state, as compared with control PCI subjects matched for age, sex, and race, and co-morbidities. While the source of these CD169^+^ cells was not definitively determined, prior work has indicated that patients with acute coronary syndrome have increased splenic metabolic activity, as measured by ^18^F-fluorodeoxyglucose–positron emission tomography, which correlates with proinflammatory remodeling of blood leukocytes and the risk of subsequent cardiovascular events.^73^ Hence, acute activation of the cardiosplenic axis after MI in humans may well include activation and mobilization of CD169^+^ MMMs to the infarct to participate in MI healing. Of note, CD169^+^ blood monocytes have been observed in humans with gastrointestinal malignancies, metastatic melanoma, and COVID-19 and were suggested to represent a heightened monocyte activation state.^74^ Moreover, splenectomy in humans has been reported as increasing the long-term risk of cardiac ischemic events and myocardial infarction.^75^ However, it is not clear whether splenectomy augments the risk of adverse post-MI remodeling in humans. Further studies will be required to further delineate this phenomenon and its implications for post-MI prognosis and outcomes.

In summary, we have established that splenic CD169^+^Tim4^+^ metallophilic marginal macrophages play a key role in the healing and repair response after MI. They comprise a key cellular component of the cardiosplenic axis both under baseline conditions and after acute myocardial injury. Splenic MMMs play a critical role in dead cell clearance, particularly of apoptotic neutrophils, the induction of reparative tissue macrophages, inflammation resolution, and proper wound healing post-MI. Expansion of the splenic marginal zone improves the LV remodeling response and alleviates inflammation, raising the possibility of splenic modulation as a therapeutic intervention during the acute MI period to prevent the late development of HF.

## ACKNOWLEDGEMENTS AND FUNDING

This work was supported by NIH grants R01 HL125735 and R01 HL157999, and VA grant I01 BX002706 to SDP; K24 HL133373 to NAL; R03 HL141620 to MX; and R01 HL137046 to TH.

